# Pilot Study to Obtain Pulmonary Endothelium from Pediatric Cardiac Catheterization

**DOI:** 10.1101/2025.09.26.25335648

**Authors:** Maxwell Mathias, Charles Sperrazza, Anas Salkini, Andrew Cave, Subhrajit Lahiri

## Abstract

Pediatric pulmonary hypertension (PH) has high morbidity and mortality, with an estimated 5-year survival of 75% regardless of underlying cause. Pediatric PH most commonly affects infants born preterm who develop lung disease of prematurity as well as those with congenital heart disease. Current therapies have been adapted from the treatment of adult PH. One barrier to the study of pediatric PH is the lack of live tissue samples for research. In this manuscript, we describe the methods to obtain pulmonary endothelium from cardiac catheterization of pediatric patients with PH. We adapted methods previously described in adult patients undergoing isolated right heart catheterization to obtain viable pulmonary endothelium from three patients. Primary cells were grown using standard endothelial culture techniques, passaged, and analyzed with immunofluorescence and flow cytometry to confirm cell type. To our knowledge, this is the first published account of pulmonary endothelial culture from pediatric patients.

## Introduction

Pediatric PH is a highly morbid condition.[1–3] Clinical trials of pediatric PH face numerous barriers, including heterogeneity of the patient population, low enrollment, and inconsistent measures of efficacy.[4] At the same time, survival of infants born extremely preterm and infants with congenital heart disease, the most common populations to develop pediatric PH, has been increasing in recent decades.[5–9] Concurrently, hospital admissions and costs of pediatric PH care have increased.[10]

Pediatric PH is defined in patients older than 3 months of age as pulmonary vascular resistance index (PVRI) of > 3.0 Woods Units x m^2^ body surface area (WU x m^2^) and mean pulmonary artery pressure (mPAP) >25 mmHg.[2,11,12] While these criteria are similar to those in adults (adult criteria were revised in 2019 at the 6^th^ World Symposium on Pulmonary Hypertension to include PVRI > 2.0 WU x m^2^ and mPAP > 20 mmHg), the epidemiology, classification, and associated comorbid conditions in pediatric patients with PH are distinct from those in adults.[11–13] The most prevalent form of PH in adults is idiopathic PH, which accounts for approximately half of all PH worldwide and exhibits a 4:1 female predominance due in part to the role of sex hormone influences on endothelial activin/inhibin signaling.[14–16] In contrast, most pediatric PH is associated with disorders of lung and cardiovascular development, and females account for only half to two-thirds of the affected population.[1,17,18] These differences in epidemiological data suggest that sex as a biological variable contributes differently to adult and pediatric PH, an important consideration in understanding pathophysiology and designing therapies for pediatric PH. In addition, endothelial dysfunction is a known aspect of pulmonary arterial hypertension in adults, but its role in pediatric PH is not clearly defined.[19,20]

While the number of available medications to treat pediatric PH has also expanded in recent decades, they have all been initially developed and tested in adults and later applied to pediatric patients, with the notable exception of inhaled nitric oxide.[21–23] Deeper understanding of the specific pathophysiology and development of novel therapies for pediatric forms of PH requires new translational tools. The following describes the isolation and initial characterization of live pulmonary endothelium from patients with pediatric PH.

## Methods

### Patient Screening and Selection

This was a prospective, convenience sample of pediatric patients aged 0 to 18 with a principal diagnosis of PH who were undergoing cardiac catheterization. Subjects and their families were approached and enrolled at their pre-procedure visit the day prior to the collection. Subjects were numbered sequentially based on when they were enrolled. The protocol was modified from a previous study that attempted, unsuccessfully, to grow endothelium from other central catheters, so the first subject in the cohort described in this manuscript was subject 17. To allow the balloon catheter to be withdrawn within the sheath at the conclusion of the case, patients were excluded if they were undergoing intervention or angiography following hemodynamics measurement.

### Collection

The legal guardians of all subjects provided informed consent. When applicable, assent was obtained from the pediatric patient.

All patients were under general anesthesia during the procedure. Routine catheterization procedures were followed including pulmonary capillary wedge pressure measurement with a 5 or 6 French Balloon Wedge Catheter (Arrow, Teleflex, USA). At the conclusion of the procedure, the balloon catheter was withdrawn into the sheath, and the sheath and catheter were removed together. The catheter tip was advanced out of the sheath into endothelial growth media (EMD Millipore SCME002) with Normocin® (InvivoGen ant-nr-1) that was warmed to 37C, cut, and transferred to a laminar flow cabinet. The catheter tip was placed into a single well of a 24-well plate that had been coated with cell attachment factor solution (Cell Applications 123-100) and incubated with media from the collection tube. The remaining media was spun down and the conditioned supernatant was added to multiple wells. The pellet was also resuspended and plated in a separate well. Half of the media from each well was transferred to a new well and the plated wells were washed every 48 hours with fresh media. Wells containing colonies were expanded into a T25 flask and further passaged and expanded to passage 3 or 4 prior to analysis.

### Analysis

Cells were washed with PBS, trypsinized, resuspended in PBS with 2% fetal bovine serum and aliquoted for flow cytometry. Samples were incubated in anti CD31-PE conjugated antibody (R&D FAB3567P), anti CD146-FITC conjugated antibody (Thermo Fisher P1H12), and anti CD45-eFluor450 conjugated antibody (Thermo Fisher HI30). Propidium Iodide was added to one aliquot for viability assessment prior to performing flow cytometry. A subculture was fixed in 10% formalin and stained for CD31 (R&D AF3628), vascular endothelial cadherin (VE-Cad, Cell Signaling D87F2), and von Willebrand Factor (vWF, Proteintech 27186-1-AP). Lastly, a subculture was incubated in DiI-conjugated acetylated LDL (DiI-acLDL, Cell Applications 022A) for 4 hours. Images were captured on an Olympus Bx43 Fluorescent microscope with a Hamamatsu ORCA-spark Digital CMOS camera in grayscale and converted to pseudocolor using FIJI/ImageJ.[24]

## Results

Patients were assigned sequential numbers in the order they were enrolled. Cultures were observed for 3 weeks after initial plating. Two of the subsequent three collections (Subject 19 and Subject 21) were successful. Subject 20 was enrolled but the case was canceled for reasons unrelated to the study. Patient demographic and cardiac catheterization data is included in Table 1 for the three successful collections. All patients underwent acute vasoreactivity testing with FiO_2_ 1.0 and inhaled nitric oxide at 20 parts per million (Table 1, Condition 2).

**Table 1:** Patient demographics, clinical data, and cardiac catheterization measurements. a. 1.2.1.f PH Associated with fetal pulmonary compression omphalocele/gastroschisis. b. 5.2.1. Inherited PH associated with *BMPR2* Mutation c. FiO_2_ 0.21 d. Blood pressures are presented as systolic/diastolic/mean in mmHg e. Pulmonary Vascular Resistance Index in Woods Units (WU) x Body surface area (BSA) = [(mmHg x min)/L] x m^2^ f. Systemic Vascular Resistance Index in WU x BSA. g. Ratio of systemic vascular resistance to pulmonary vascular resistance h. Cardiac index (Cardiac Output x BSA) in L/min/m^2^; for subject 17, Qp (pulmonary) and Qs (systemic) indices are included i. FiO_2_ 1.0 and inhaled nitric oxide 20 parts per million

| Subject # | 17 | 19 | 21 |
| --- | --- | --- | --- |
| Age (years) | < 5 | 11-15 | 6-10 |
| Sex | F | M | M |
| Weight | 6.0 kg | 65.6kg | 29.0kg |
| Primary Diagnosis (Panama 2011)<sup>11</sup> | 1.2.1.f.<sup>a</sup> | 5.2.1.<sup>b</sup> | 5.2.1.<sup>b</sup> |
| Other Diagnoses | Anterior malalignment VSD | Autism Spectrum Disorder | Mild Obstructive Sleep Apnea |
| Medications | Tadalafil, furosemide, chlorthiazide, spironolactone | Tadalafil, bosentan | Sildenafil, macicentan |
| **Condition 1<sup>c</sup>** |  |  |  |
| Pulmonary Artery | 59/21/39<sup>d</sup> | 54/11/36 | 74/25/41 |
| Aorta | 79/55/66 | 82/48/59 | 99/48/64 |
| Right Atrium | 7/5/5 | 8/4/6 | 14/10/12 |
| Pulmonary Capillary Wedge Pressure | 11 | 10 | 15 |
| PVRI<sup>e</sup> | 3.8 | 5.2 | 6.5 |
| SVRI<sup>f</sup> | 23.4 | 9.8 | 9.2 |
| PVR/SVR<sup>g</sup> | 0.16 | 0.53 | 0.71 |
| Cardiac Index<sup>h</sup> | Qp = 7.8; Qs = 2.6 | 5.6 | 5.9 |
| **Condition 2<sup>i</sup>** |  |  |  |
| Pulmonary Artery | 48/19/33 | 59/7/38 | 74/32/49 |
| Aorta | 75/50/64 | 95/49/63 | 103/51/68 |
| Right Atrium | 8/5/5 | 11/6/7 | 15/11/12 |
| Pulmonary Capillary Wedge Pressure | 11 | 11 | 16 |
| PVRI | 1.6 | 2.3 | 5.9 |
| SVRI | 16.9 | 4.9 | 10 |
| PVR/SVR | 0.10 | 0.47 | 0.59 |
| Cardiac Index | Qp = 13.2; Qs = 3.5 | 11.5 | 5.7 |

For successful collections, endothelial cells were first visible 2-10 days after collection and were confluent within 2-3 weeks. In all three samples assessed, more than 98% of cells were CD31+/CD146+/CD45- and 85-95% viable by propidium iodide staining (Figure 1). Fluorochrome controls and gating parameters can be found in supplementary data (Figures S1-S3). Fixed cells stained positive for CD31, VE-Cad, and vWF (Figs. 2, S4, S5). Lastly, cells demonstrated uptake of DiI-conjugated ac-LDL (Figs. 2, S4, S5).

**Figure 1:**
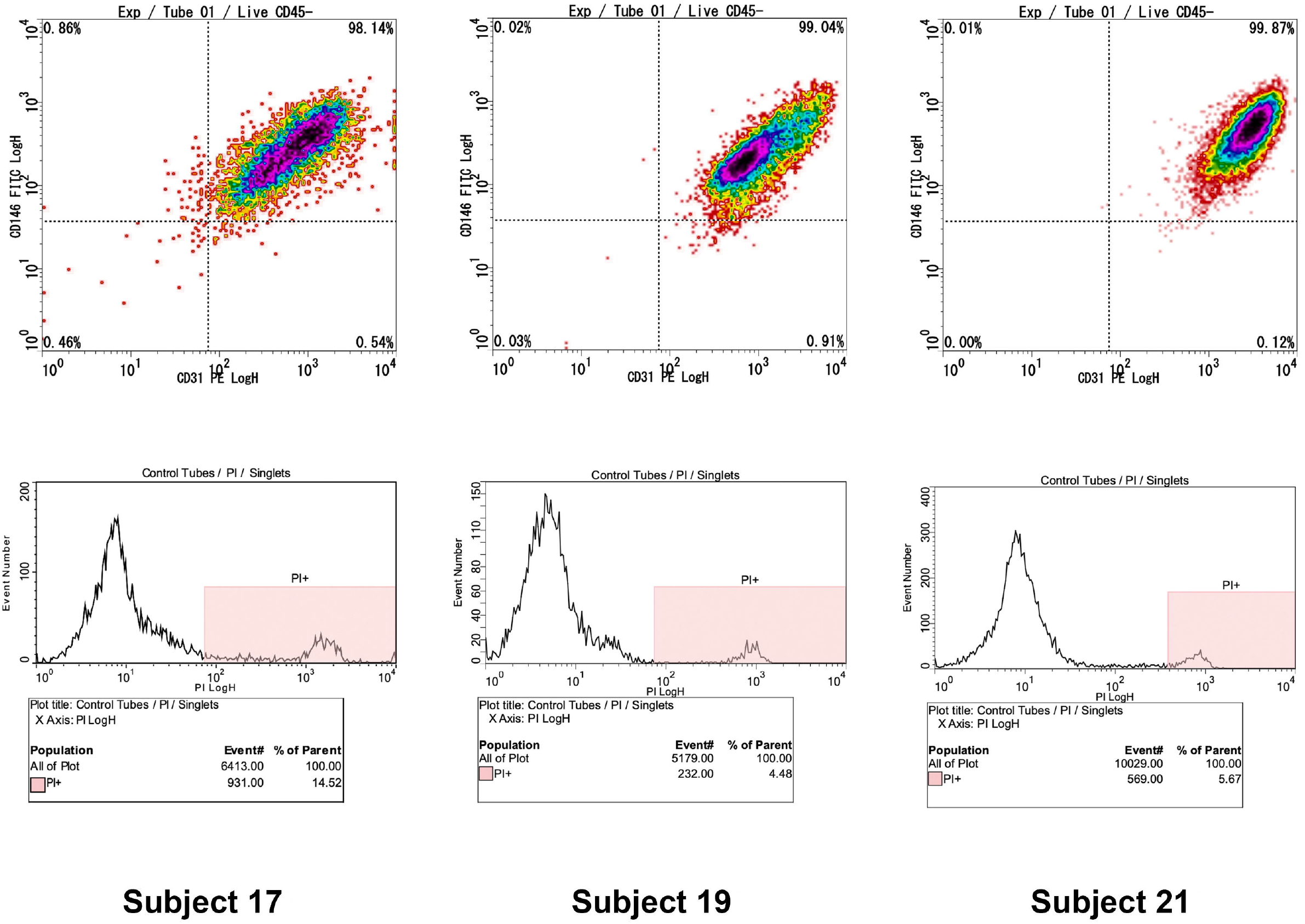
Flow cytometry data from Subjects 17, 19, and 21. Top panel shows flow cytometry data with CD31 positivity on x-axis and CD146 positivity on y-axis. All cells included were CD45 negative. 10,000 cells were counted for each subject. Bottom panel shows propidium iodide viability staining. At least 5,000 cells were assessed for viability which ranged from 85-95%.

**Figure 2:**
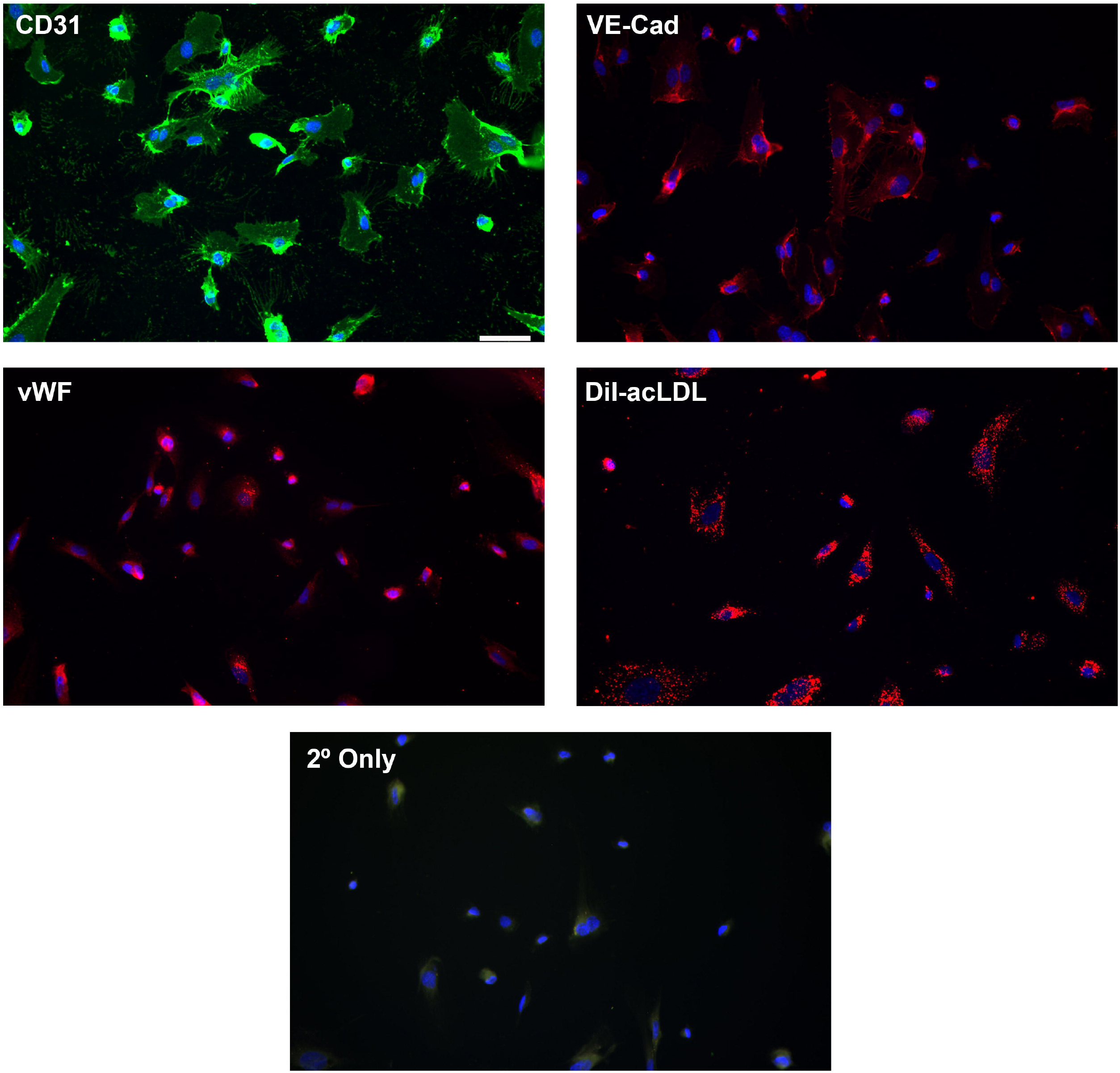
Representative images of formalin fixed cells from subject 21 staining for endothelial markers CD31, vascular endothelial cadherin (VE-Cad), von Willebrand Factor (vWF), and 4-hour uptake of DiI-conjugated acetylated LDL (DiI-acLDL). All images obtained at 20x magnification, scale bar in CD31 image is 50μm. Negative control with secondary antibodies only is shown at the bottom.

## Discussion

The present study describes methods adapted from Ventetuolo et al. to isolate pulmonary endothelium from pediatric patients undergoing cardiac catheterization.[25] Cells grown from catheter balloon tips in three pediatric patients were viable, exhibited multiple markers of endothelial lineage, did not express CD45, a marker of hematopoietic lineage, and exhibited uptake of ac-LDL a feature found only in endothelium and macrophages.

Knowledge of pulmonary endothelial development, cell specialization, and cell-cell interactions in pediatric lung diseases has expanded rapidly in the era of single cell RNA sequencing.[26,27] However, these studies of pulmonary endothelium have relied on lung explants, post-mortem tissue, or animal models.[28–31] Additional translational tools to uncover mechanisms, evolution, and therapeutic response to pediatric PH are needed. Further collection and deeper molecular phenotyping of catheter-derived pulmonary endothelium may serve as one such tool.

Pollett et al first described the isolation of viable pulmonary endothelium in 2013 from catheter balloon tips in a cohort of adult patients undergoing right heart catheterization.[32] Since then, his group and others have refined these methods and applied them to larger cohorts.[25,33,34] Singh et al describe a detailed analysis of the largest published cohort to date, a study of 54 samples from 49 patients.[35] In this study, a comparison of bulk RNA sequencing in patients with PH and controls found differential expression in genes associated with Wnt and Bone Morphogenetic Protein (BMP) signaling that was consistent across PH diagnostic categories and persisted through early passaging of cells.[35] Dysregulated endothelial BMP signaling is a well-established aspect of idiopathic PH, and downstream effectors of BMP signaling are targeted by the recently approved drug sotatercept, which alters endothelial and smooth muscle cell proliferation and has shown remarkable efficacy in severe PH.[36–39] The efficacy of sotatercept supports the role of endothelial dysfunction in PH pathogenesis and illustrates the potential for isolated pulmonary endothelium from patients with PH as a translational resource. Future applications of these collections could include individualized testing of novel therapeutics, which may have distinct effects on the growing and developing lungs and vasculature of pediatric patients. In addition, serial collections in patients who undergo repeat catheterizations may provide cytopathologic correlate to disease progression or remission.

While cardiac catheterization is a recommended part of evaluation for pediatric patients with PH, there are risks unique to this population.[12,40] In contrast to right heart catheterization in adults, most children require general anesthesia for cardiac catheterization. Like adults, the risk of complications from anesthesia are significantly greater in patients with PH, though these can be mitigated through multidisciplinary collaboration between pediatric cardiac anesthesia, pediatric cardiologists specializing in PH, and pediatric interventional cardiologists.[41–44] Thus, guidelines from the American Heart Association and American Thoracic Society recommend that catheterization occur only at centers experienced in management of pediatric PH to limit the associated morbidity and mortality.[12,41]

To our knowledge, this is the first report of isolation of viable pulmonary endothelium from a balloon wedge catheter in pediatric patients. Further isolation and analysis of pulmonary endothelium from pediatric patients could provide insight into the pathophysiology of pediatric PH and serve as a preclinical resource for testing therapies.

## Data Availability

All data produced in the present study are available upon reasonable request to the authors

## Author Contributions

M.M., C.S., A.S., and S.L. designed and implemented the study. M.M. processed and analyzed the cells and drafted the initial manuscript. All authors were involved in critical revisions of the manuscript. All authors read and approve of the final manuscript.

## Acknowledgements

We thank Dr. Corey Ventetuolo and Mandy Pereira for guidance in methodology. We also thank the Institutional Research Core Facility at the University of Oklahoma Health School of Medicine for use of the Flow Cytometry and Imaging facility.

## Ethics Statement

This study was approved by the institutional review board at the University of Oklahoma Health Sciences Center (IRB # 15767).

## Guarantor

M.M. is the guarantor of the manuscript and takes full responsibility for the integrity of the research described.

