## Supplemental Figure 1 for "Pilot Study to Obtain Pulmonary Endothelium from Pediatric Cardiac Catheterization"

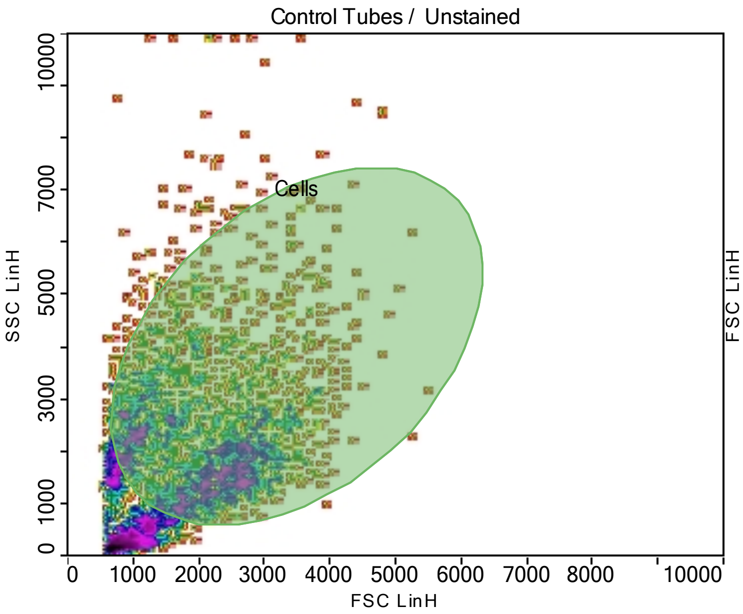


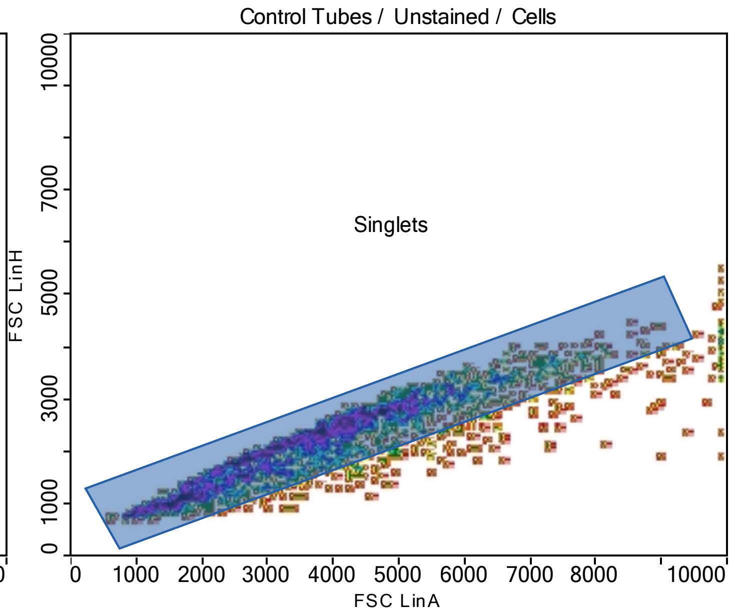


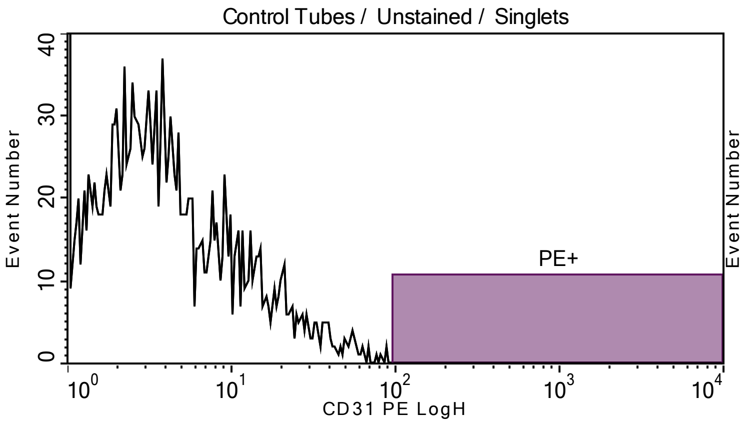


Plot title: Control Tubes / Unstained / Singlets
X Axis: CD31 PE LogH

| Population | Event# | % of Parent |
|---|---|---|
| All of Plot | 1978.00 | 100.00 |
| PE+ | 0.00 | 0.00 |

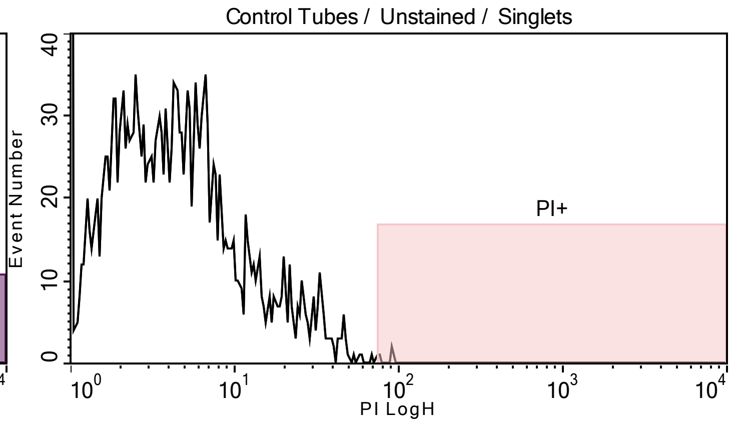


Plot title: Control Tubes / Unstained / Singlets
X Axis: PI LogH

| Population | Event# | % of Parent |
|---|---|---|
| All of Plot | 1978.00 | 100.00 |
| PI+ | 3.00 | 0.15 |

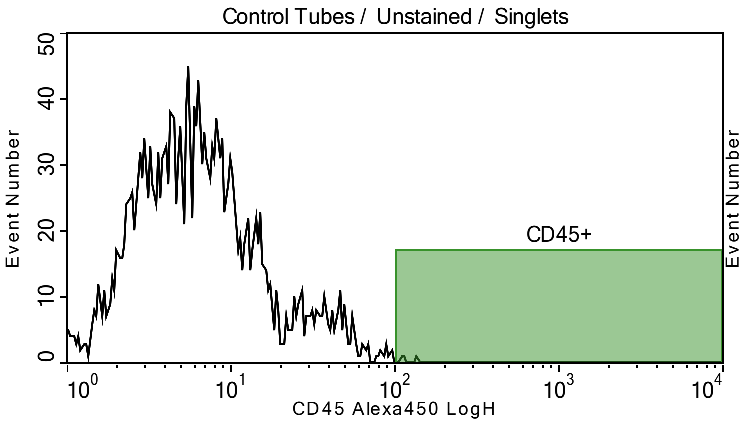


Plot title: Control Tubes / Unstained / Singlets
X Axis: CD45 Alexa450 LogH

| Population | Event# | % of Parent |
|---|---|---|
| All of Plot | 1978.00 | 100.00 |
| CD45+ | 3.00 | 0.15 |

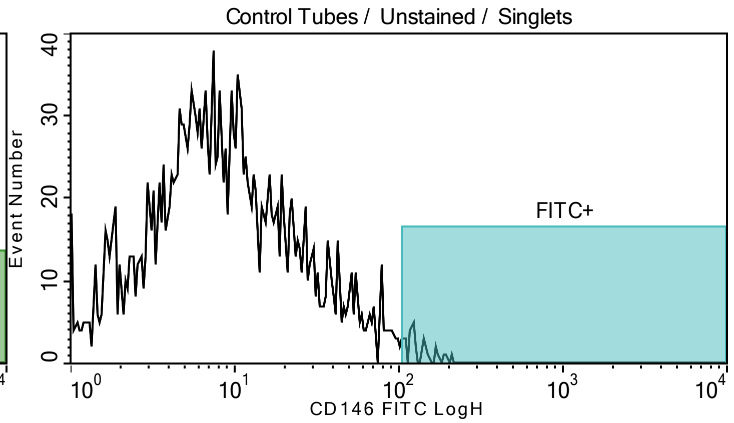


Plot title: Control Tubes / Unstained / Singlets
X Axis: CD146 FITC LogH

| Population | Event# | % of Parent |
|---|---|---|
| All of Plot | 1978.00 | 100.00 |
| FITC+ | 28.00 | 1.42 |

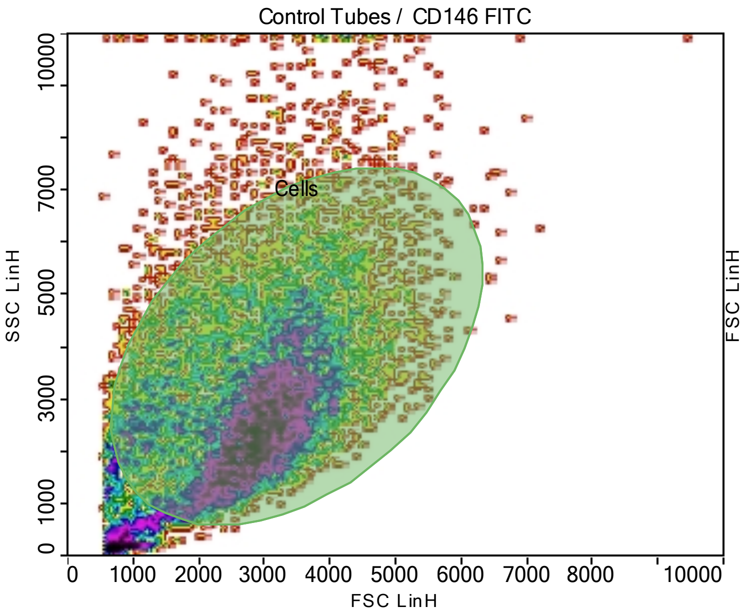


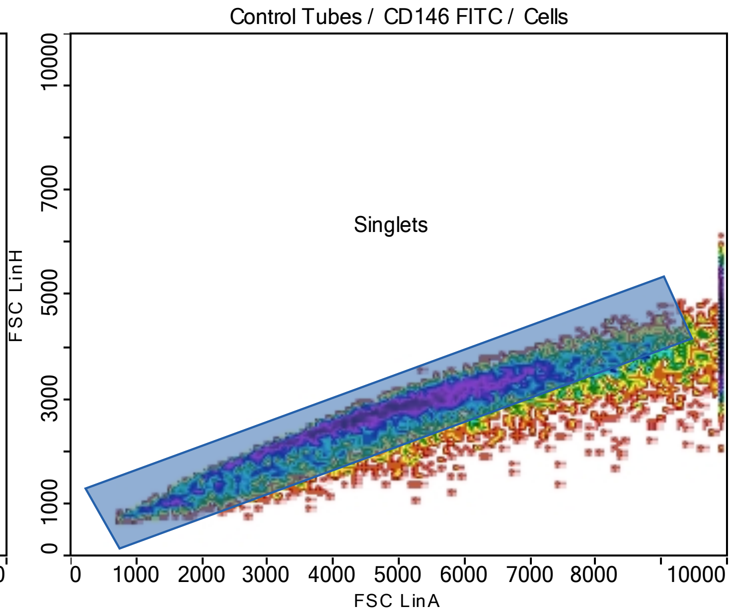


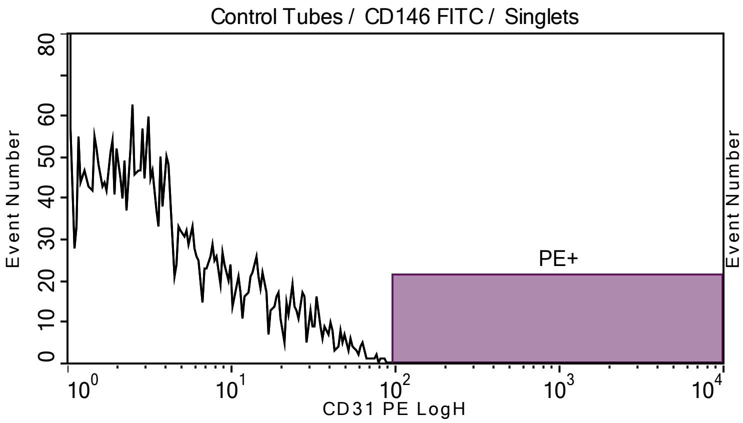


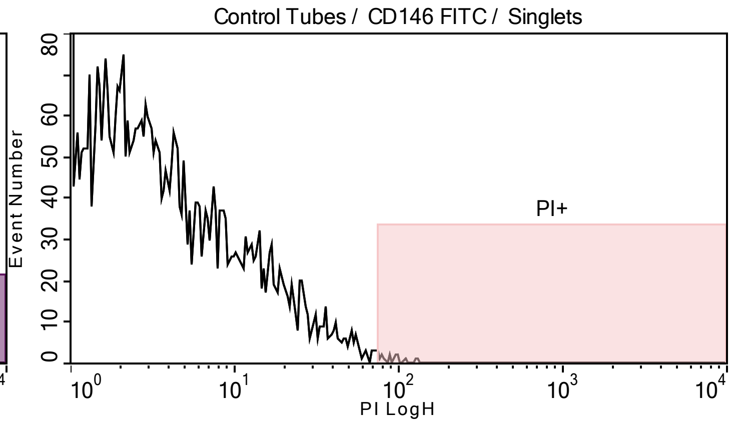


Plot title: Control Tubes / CD146 FITC / Singlets
X Axis: CD31 PE LogH

| Population | Event# | % of Parent |
|---|---|---|
| All of Plot | 7132.00 | 100.00 |
| PE+ | 0.00 | 0.00 |

Plot title: Control Tubes / CD146 FITC / Singlets
X Axis: PI LogH

| Population | Event# | % of Parent |
|---|---|---|
| All of Plot | 7132.00 | 100.00 |
| PI+ | 15.00 | 0.21 |

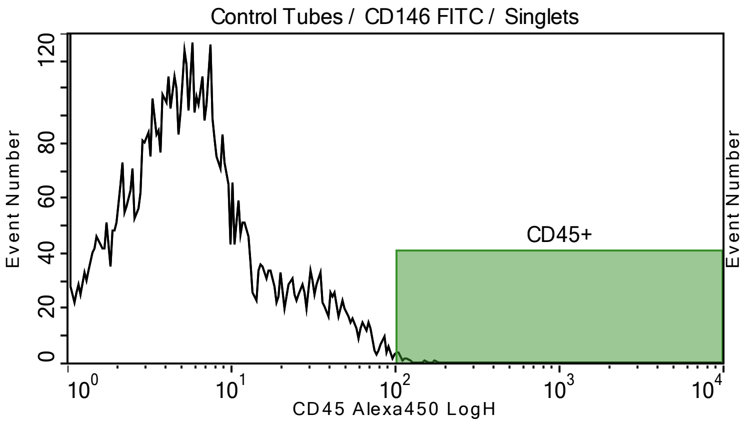


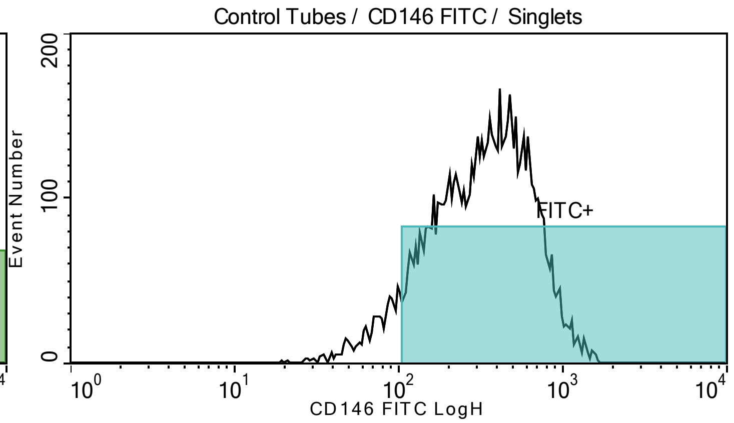


Plot title: Control Tubes / CD146 FITC / Singlets
X Axis: CD45 Alexa450 LogH

| Population | Event# | % of Parent |
|---|---|---|
| All of Plot | 7132.00 | 100.00 |
| CD45+ | 16.00 | 0.22 |

Plot title: Control Tubes / CD146 FITC / Singlets
X Axis: CD146 FITC LogH

| Population | Event# | % of Parent |
|---|---|---|
| All of Plot | 7132.00 | 100.00 |
| FITC+ | 6519.00 | 91.40 |

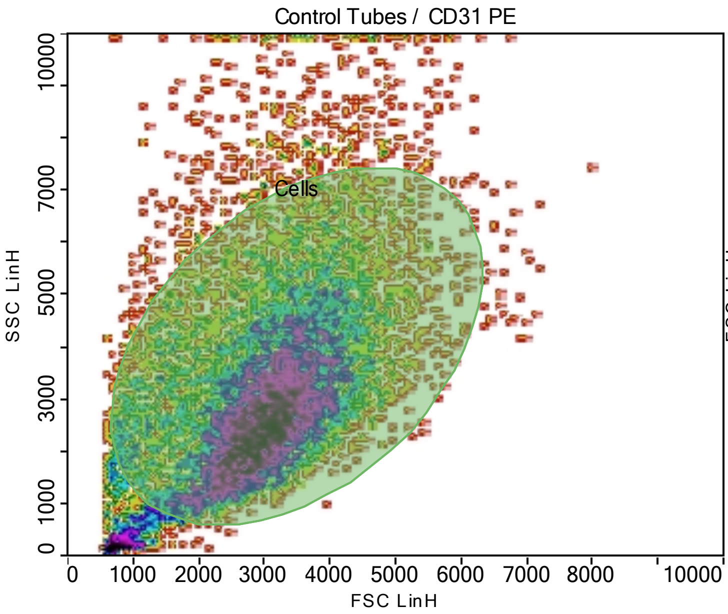


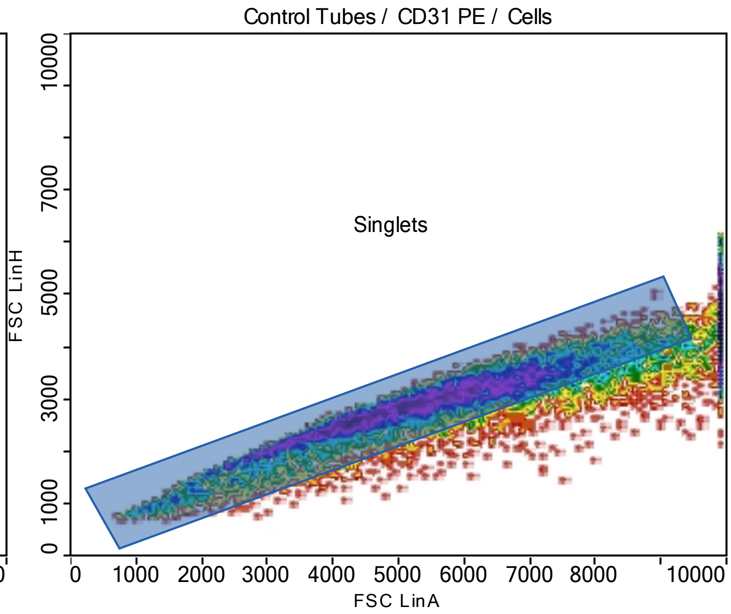


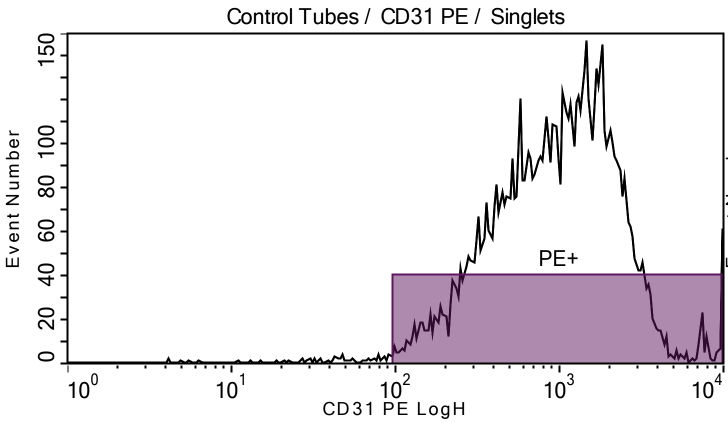


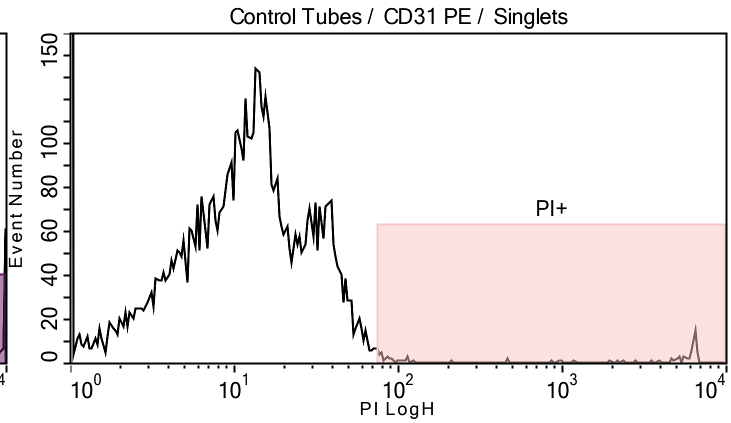


Plot title: Control Tubes / CD31 PE / Singlets
X Axis: CD31 PE LogH

| **Population** | **Event#** | **% of Parent** |
|---|---|---|
| All of Plot | 7078.00 | 100.00 |
| PE+ | 7016.00 | 99.12 |

Plot title: Control Tubes / CD31 PE / Singlets
X Axis: PI LogH

| **Population** | **Event#** | **% of Parent** |
|---|---|---|
| All of Plot | 7078.00 | 100.00 |
| PI+ | 80.00 | 1.13 |

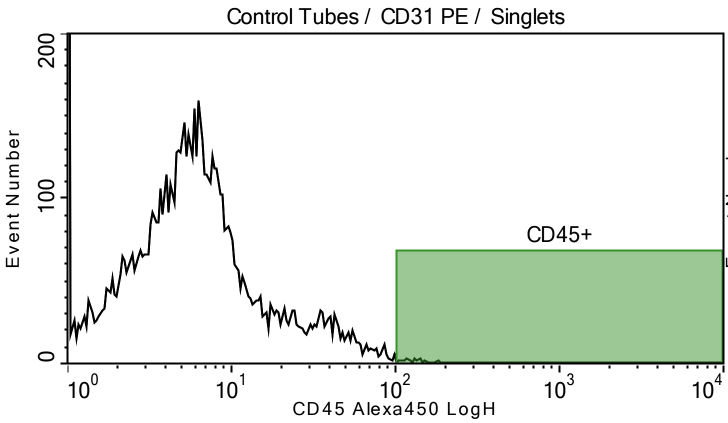


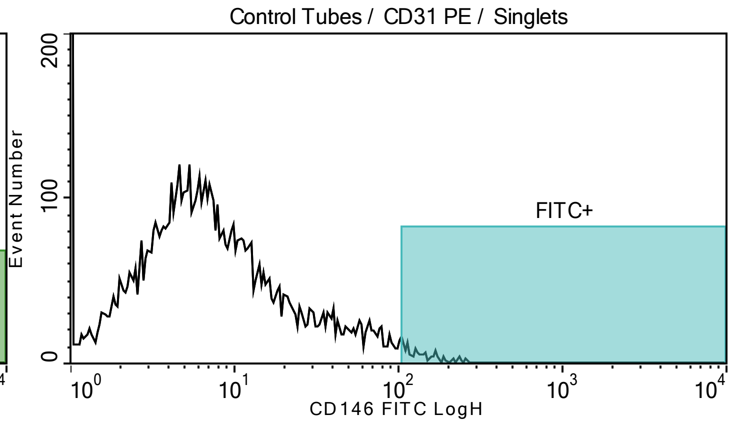


Plot title: Control Tubes / CD31 PE / Singlets
X Axis: CD45 Alexa450 LogH

| **Population** | **Event#** | **% of Parent** |
|---|---|---|
| All of Plot | 7078.00 | 100.00 |
| CD45+ | 27.00 | 0.38 |

Plot title: Control Tubes / CD31 PE / Singlets
X Axis: CD146 FITC LogH

| **Population** | **Event#** | **% of Parent** |
|---|---|---|
| All of Plot | 7078.00 | 100.00 |
| FITC+ | 132.00 | 1.86 |

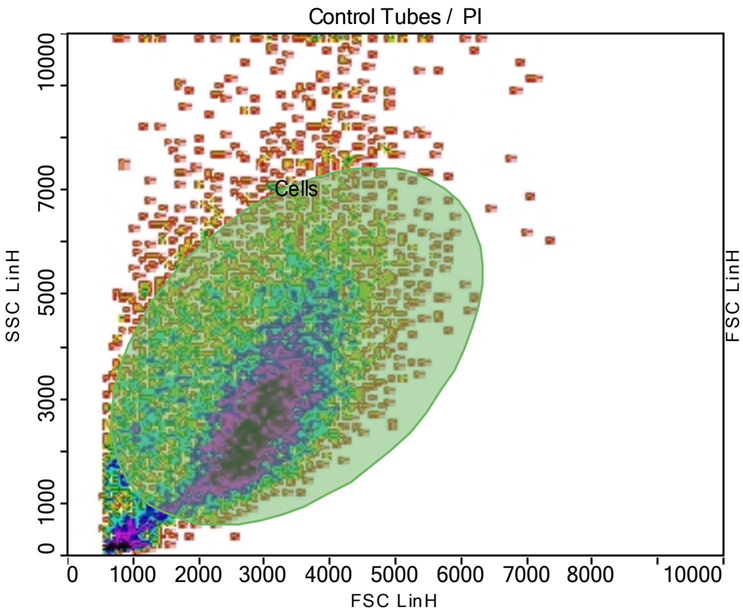


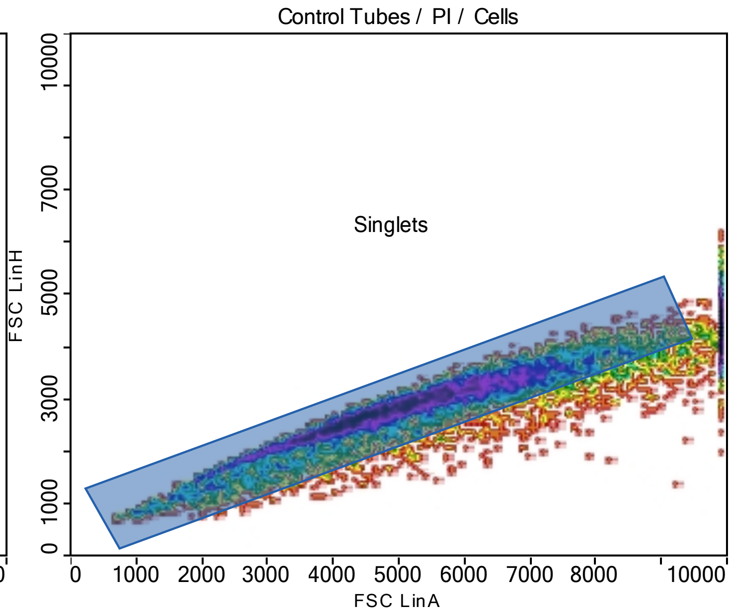


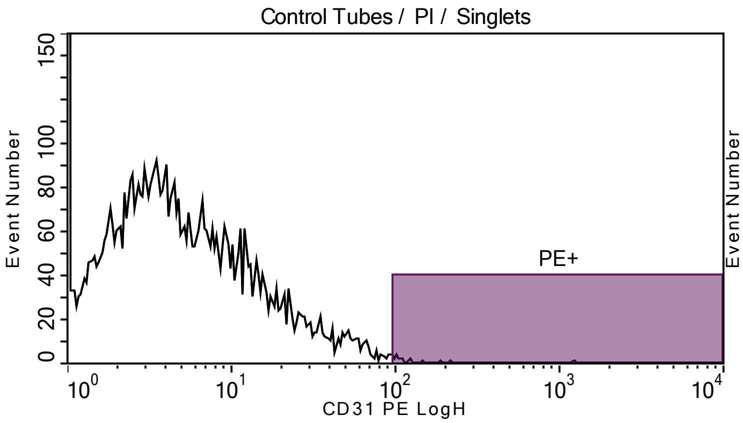


Plot title: Control Tubes / PI / Singlets
X Axis: CD31 PE LogH

| Population | Event# | % of Parent |
|---|---|---|
| All of Plot | 6413.00 | 100.00 |
| PE+ | 16.00 | 0.25 |

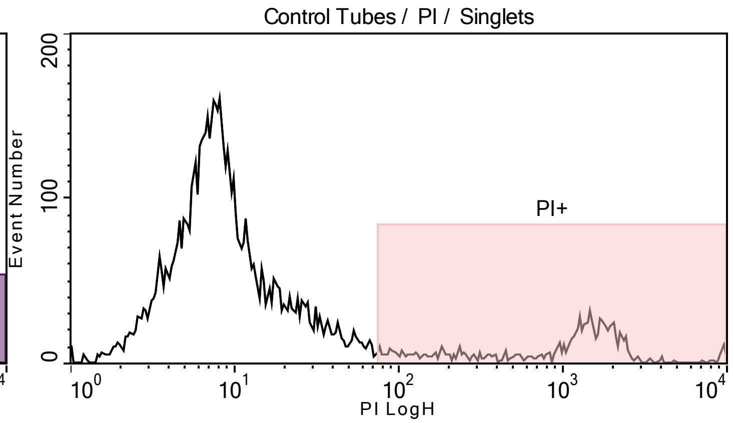


Plot title: Control Tubes / PI / Singlets
X Axis: PI LogH

| Population | Event# | % of Parent |
|---|---|---|
| All of Plot | 6413.00 | 100.00 |
| PI+ | 931.00 | 14.52 |

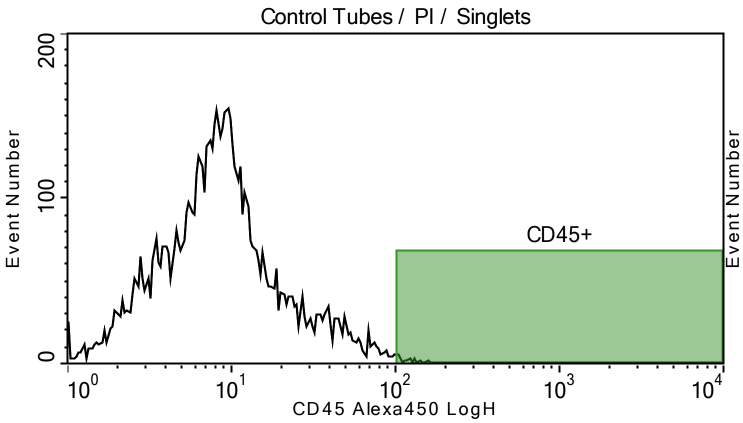


Plot title: Control Tubes / PI / Singlets
X Axis: CD45 Alexa450 LogH

| Population | Event# | % of Parent |
|---|---|---|
| All of Plot | 6413.00 | 100.00 |
| CD45+ | 32.00 | 0.50 |

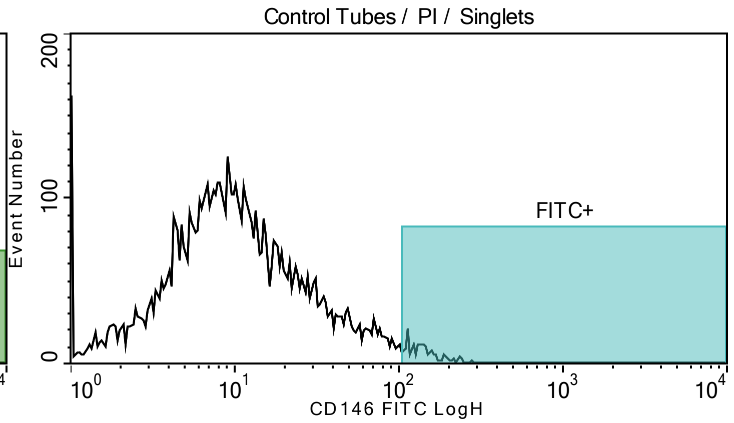


Plot title: Control Tubes / PI / Singlets
X Axis: CD146 FITC LogH

| Population | Event# | % of Parent |
|---|---|---|
| All of Plot | 6413.00 | 100.00 |
| FITC+ | 161.00 | 2.51 |

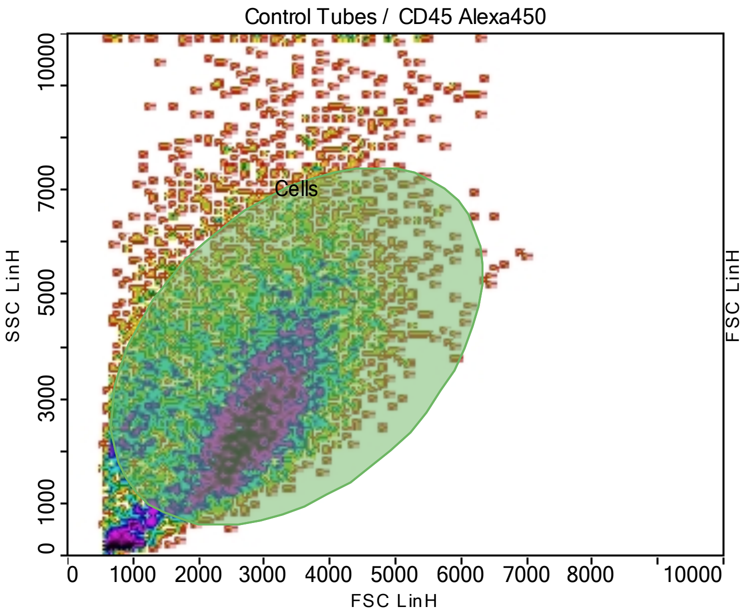


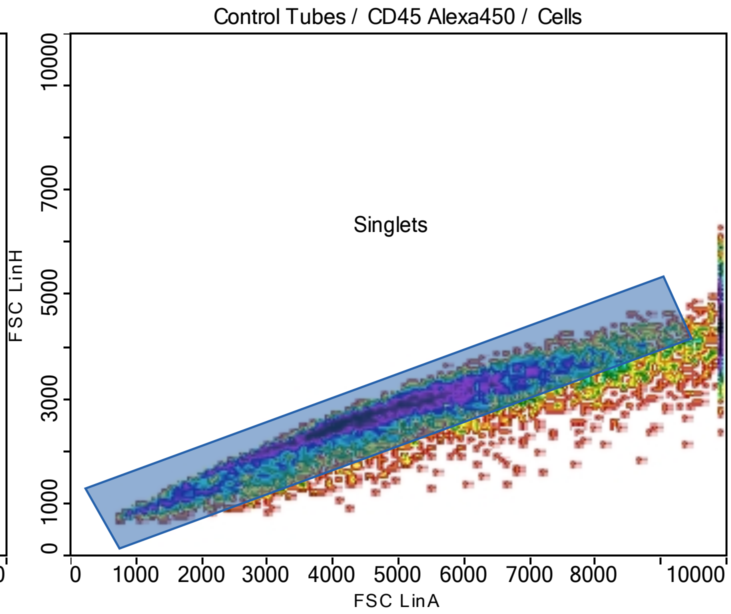


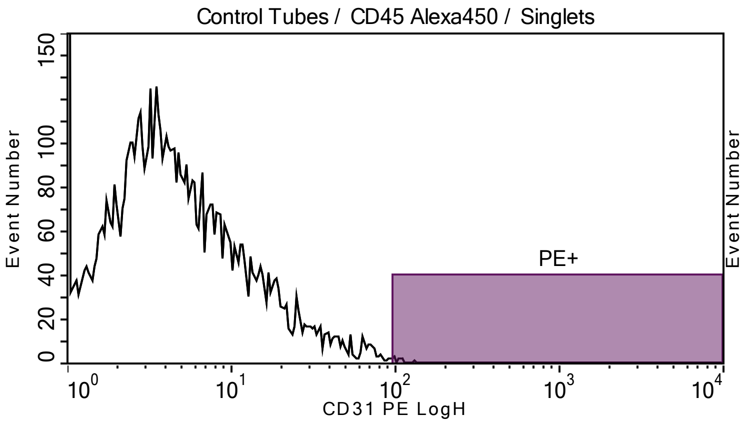


Plot title: Control Tubes / CD45 Alexa450 / Singlets
X Axis: CD31 PE LogH

| Population | Event# | % of Parent |
|---|---|---|
| All of Plot | 6545.00 | 100.00 |
| PE+ | 8.00 | 0.12 |

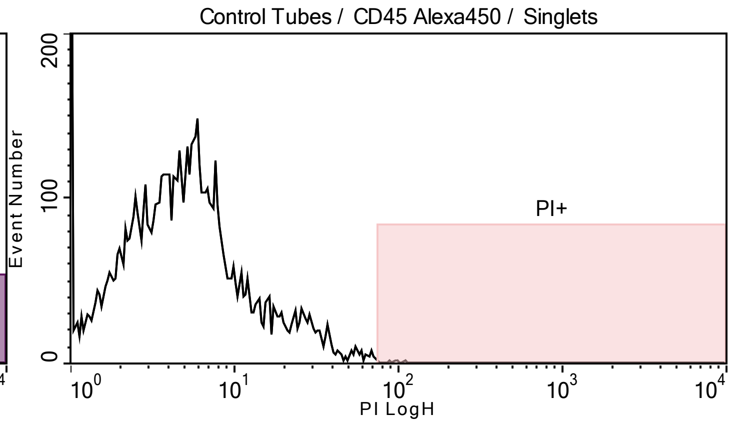


Plot title: Control Tubes / CD45 Alexa450 / Singlets
X Axis: PI LogH

| Population | Event# | % of Parent |
|---|---|---|
| All of Plot | 6545.00 | 100.00 |
| PI+ | 17.00 | 0.26 |

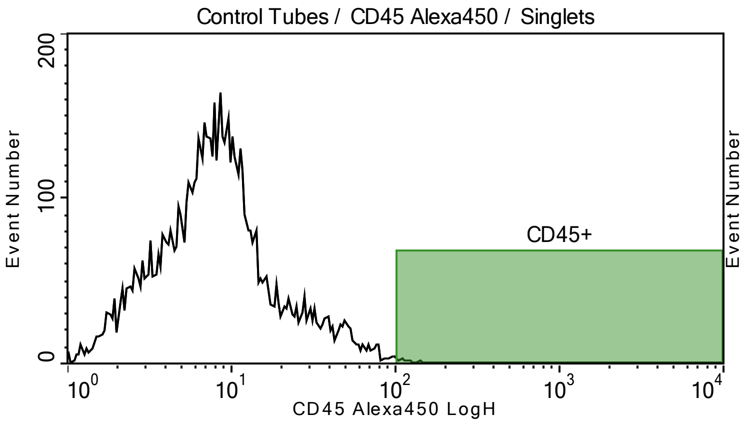


Plot title: Control Tubes / CD45 Alexa450 / Singlets
X Axis: CD45 Alexa450 LogH

| Population | Event# | % of Parent |
|---|---|---|
| All of Plot | 6545.00 | 100.00 |
| CD45+ | 20.00 | 0.31 |

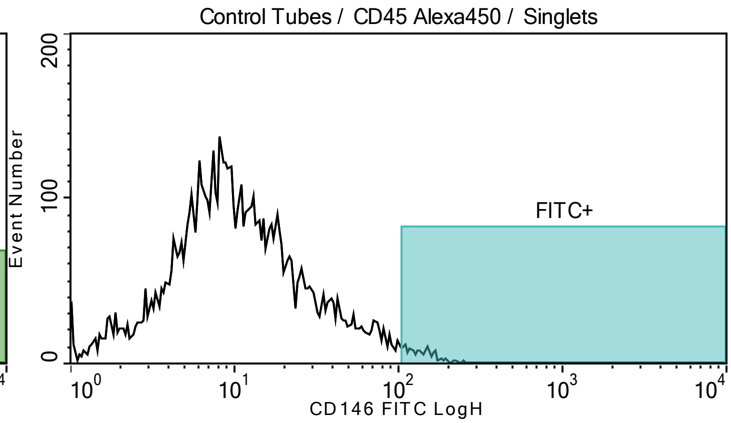


Plot title: Control Tubes / CD45 Alexa450 / Singlets
X Axis: CD146 FITC LogH

| Population | Event# | % of Parent |
|---|---|---|
| All of Plot | 6545.00 | 100.00 |
| FITC+ | 128.00 | 1.96 |

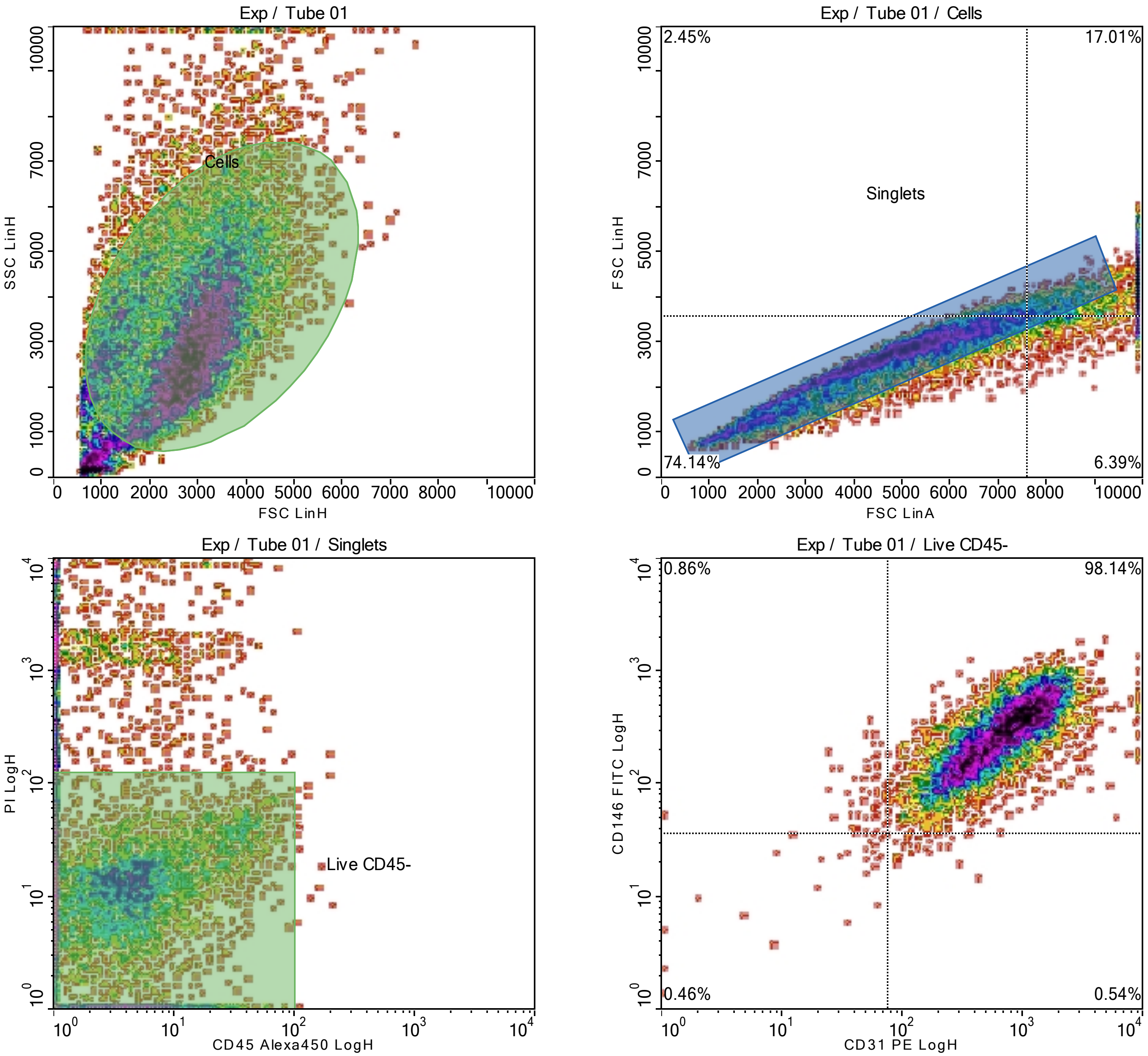

Exp / Tube 01
Cells
SSC LinH
FSC LinH
Exp / Tube 01 / Cells
2.45%
17.01%
Singlets
74.14%
6.39%
FSC LinH
FSC LinA
Exp / Tube 01 / Singlets
Live CD45-
PI LogH
CD45 Alexa450 LogH
Exp / Tube 01 / Live CD45-
0.86%
98.14%
0.46%
0.54%
CD146 FITC LogH
CD31 PE LogH
