## Supplemental Figure 2 for "Pilot Study to Obtain Pulmonary Endothelium from Pediatric Cardiac Catheterization"

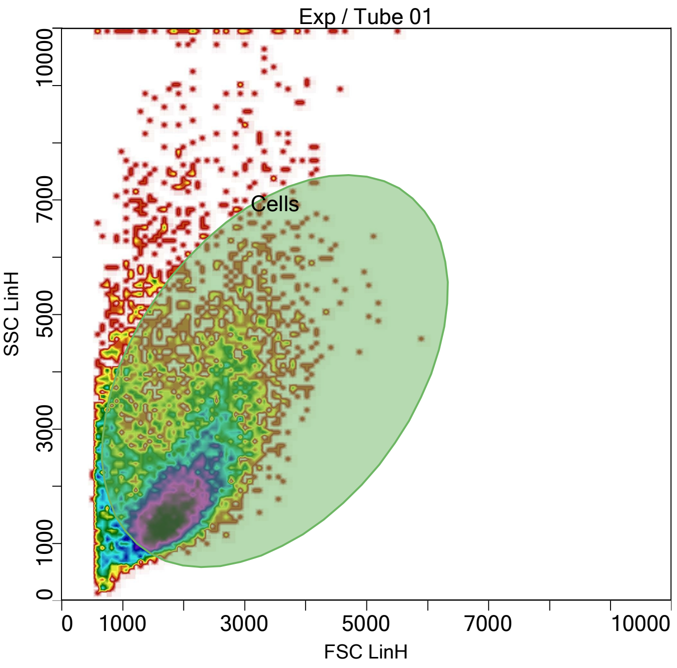


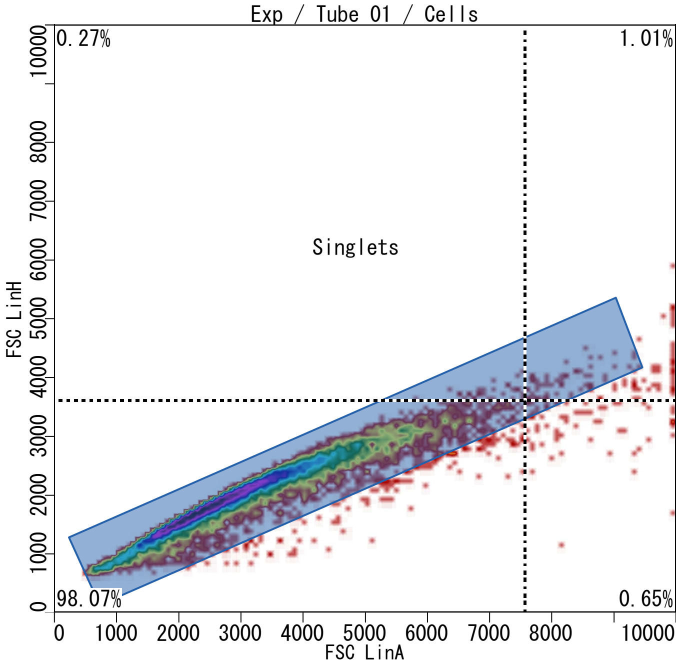


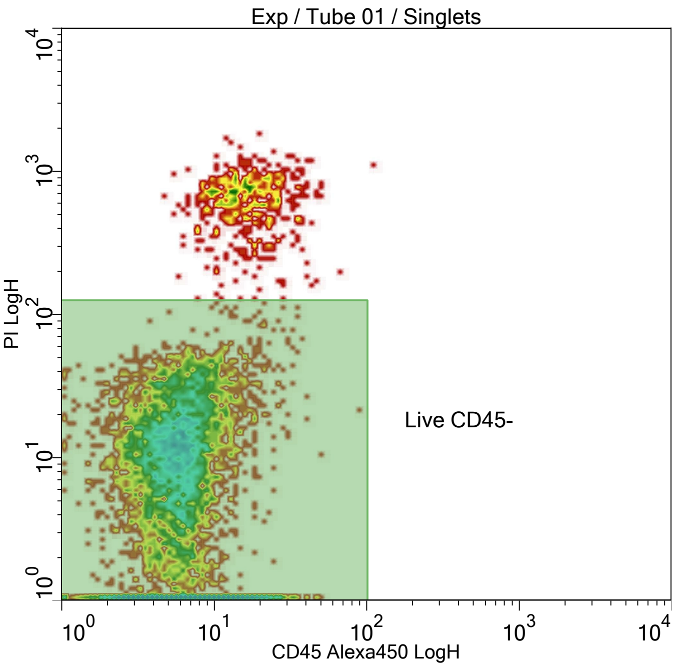


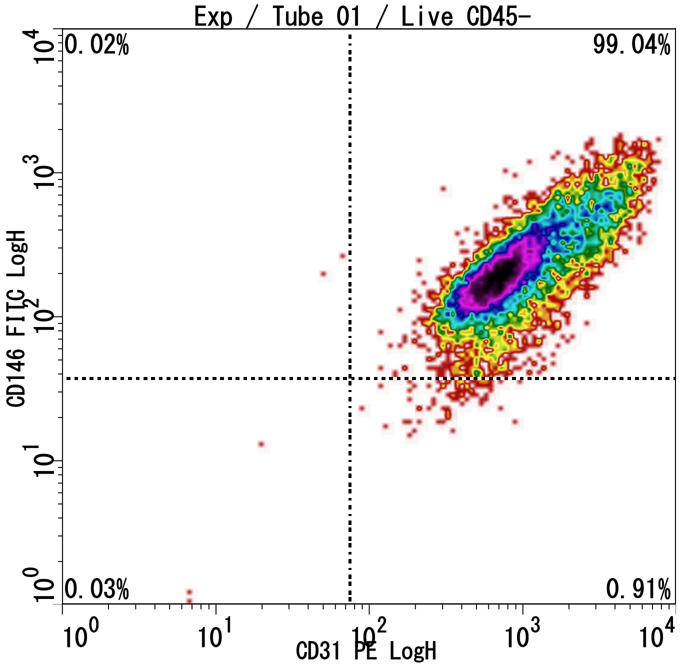


Cytometry Without Compromise
www.stratedigm.com

Fitc PE PI a450 7-21-25 #19, printed at 7/21/2025 14:49:26
Printed by CellCapTure v4.1 RC27, Copyright 2022 Stratedigm
Licensed to: Oklahoma University OUHSC

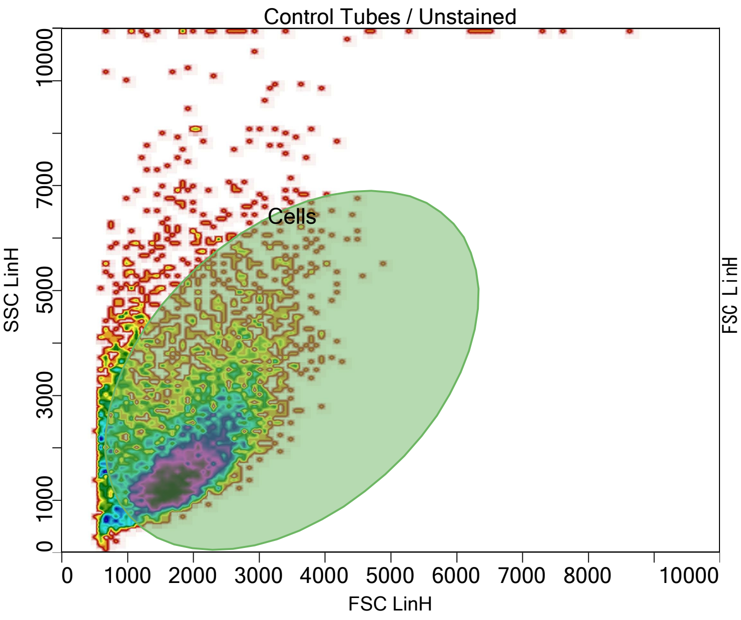


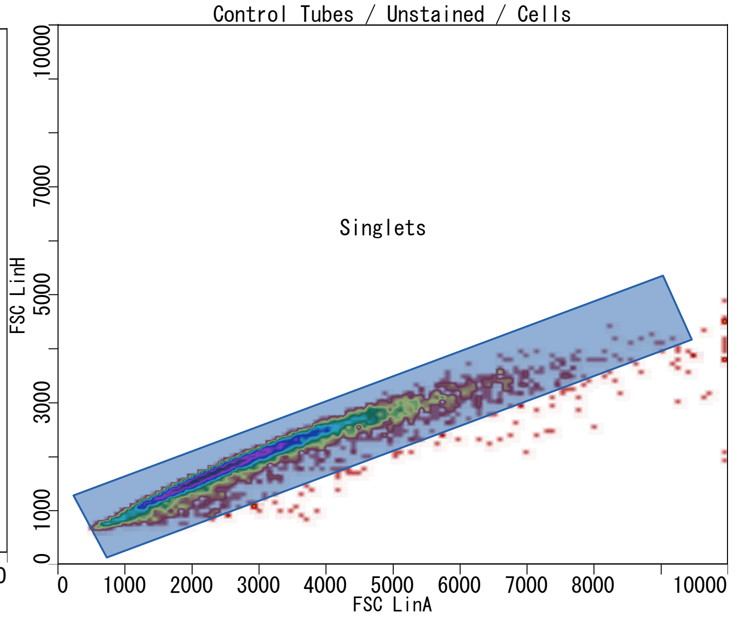


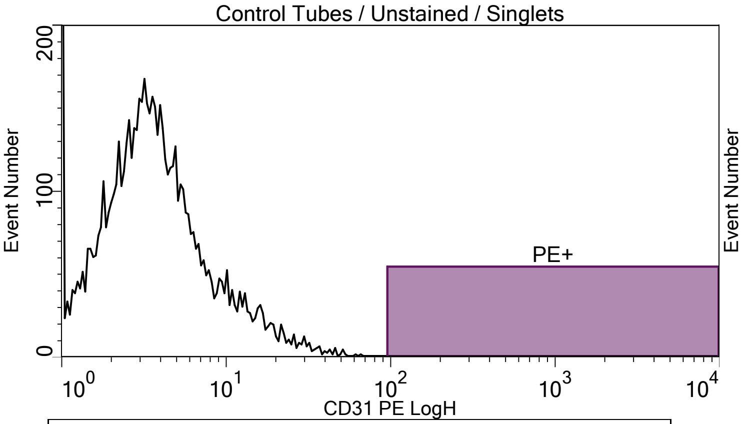


Plot title: Control Tubes / Unstained / Singlets
X Axis: CD31 PE LogH

| Population | Event# | % of Parent |
|---|---|---|
| All of Plot | 6674.00 | 100.00 |
| PE+ | 0.00 | 0.00 |

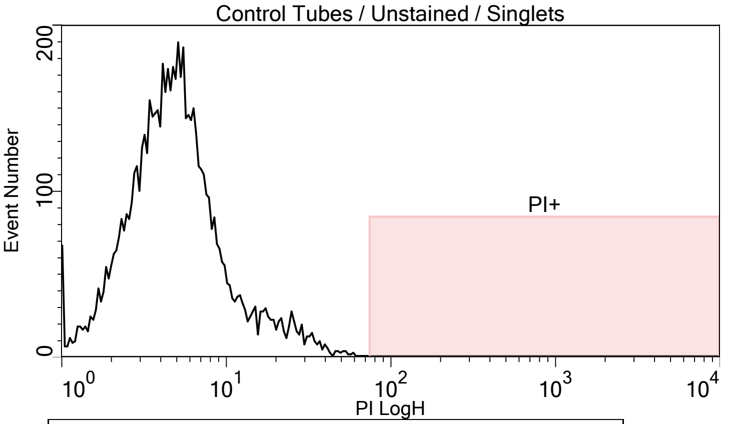


Plot title: Control Tubes / Unstained / Singlets
X Axis: PI LogH

| Population | Event# | % of Parent |
|---|---|---|
| All of Plot | 6674.00 | 100.00 |
| PI+ | 0.00 | 0.00 |

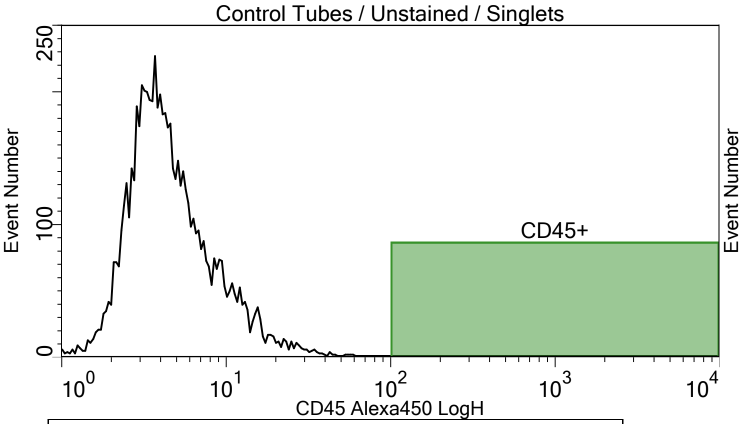


Plot title: Control Tubes / Unstained / Singlets
X Axis: CD45 Alexa450 LogH

| Population | Event# | % of Parent |
|---|---|---|
| All of Plot | 6674.00 | 100.00 |
| CD45+ | 0.00 | 0.00 |

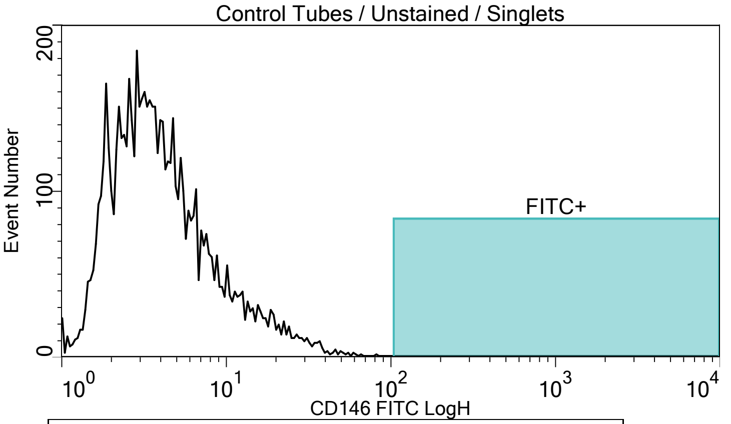


Plot title: Control Tubes / Unstained / Singlets
X Axis: CD146 FITC LogH

| Population | Event# | % of Parent |
|---|---|---|
| All of Plot | 6674.00 | 100.00 |
| FITC+ | 0.00 | 0.00 |

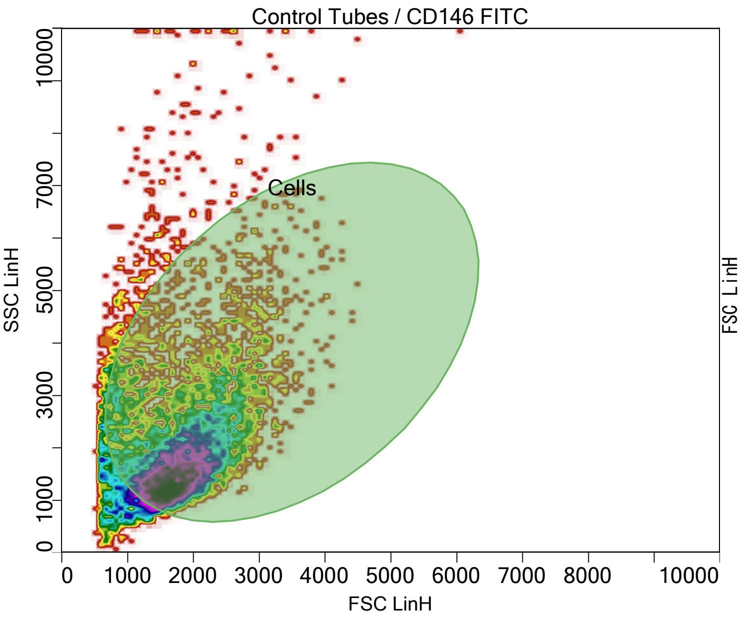


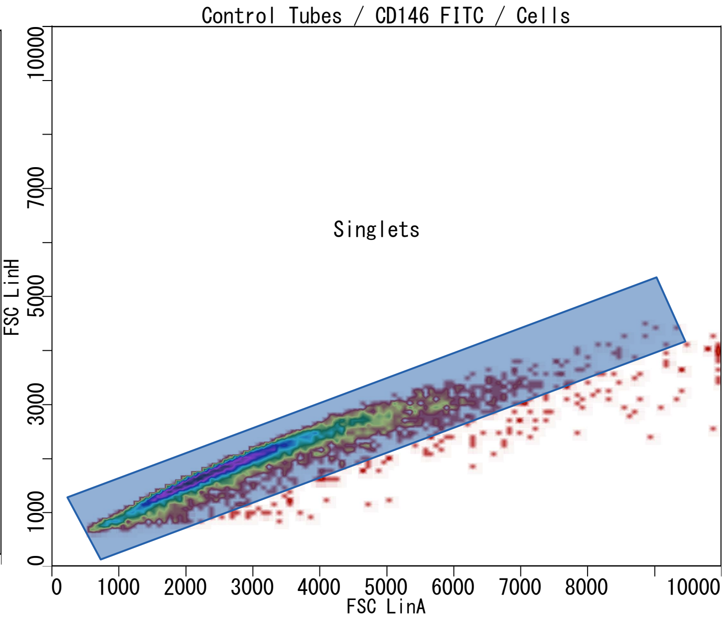


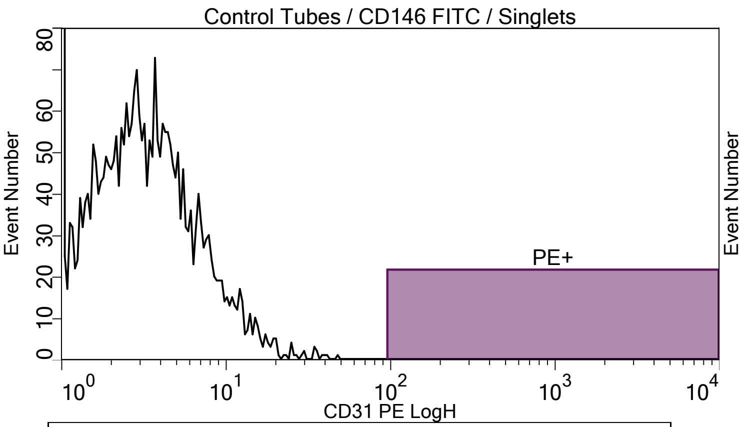


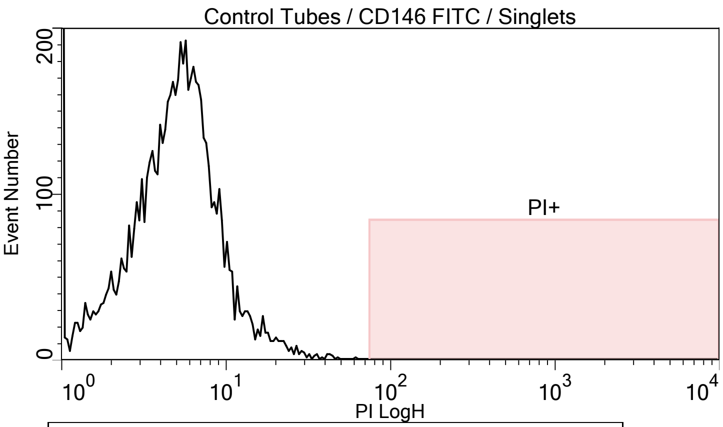


Plot title: Control Tubes / CD146 FITC / Singlets
X Axis: CD31 PE LogH

| Population | Event# | % of Parent |
|---|---|---|
| All of Plot | 6820.00 | 100.00 |
| PE+ | 0.00 | 0.00 |

Plot title: Control Tubes / CD146 FITC / Singlets
X Axis: PI LogH

| Population | Event# | % of Parent |
|---|---|---|
| All of Plot | 6820.00 | 100.00 |
| PI+ | 0.00 | 0.00 |

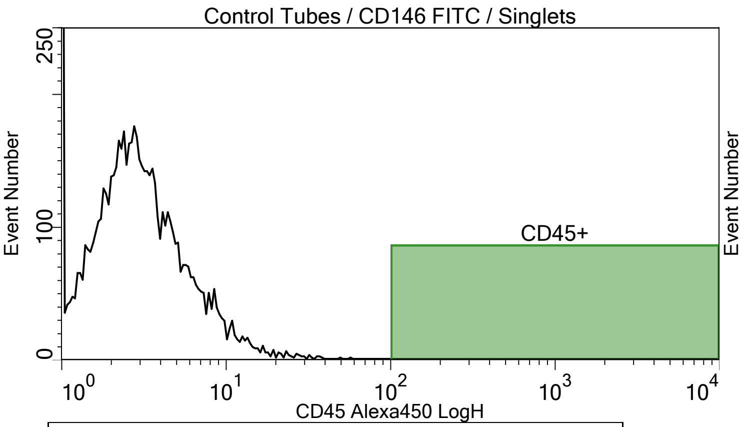


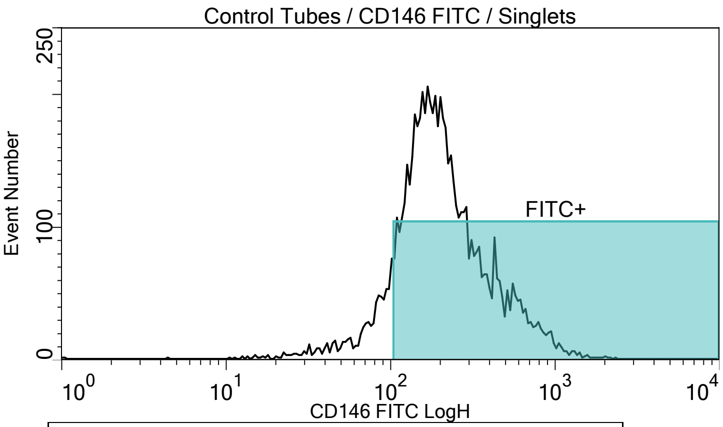


Plot title: Control Tubes / CD146 FITC / Singlets
X Axis: CD45 Alexa450 LogH

| Population | Event# | % of Parent |
|---|---|---|
| All of Plot | 6820.00 | 100.00 |
| CD45+ | 0.00 | 0.00 |

Plot title: Control Tubes / CD146 FITC / Singlets
X Axis: CD146 FITC LogH

| Population | Event# | % of Parent |
|---|---|---|
| All of Plot | 6820.00 | 100.00 |
| FITC+ | 6044.00 | 88.62 |

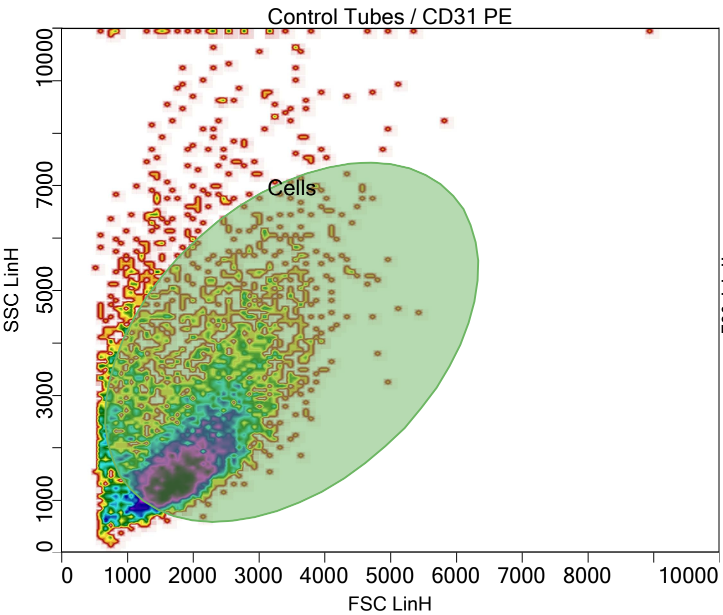


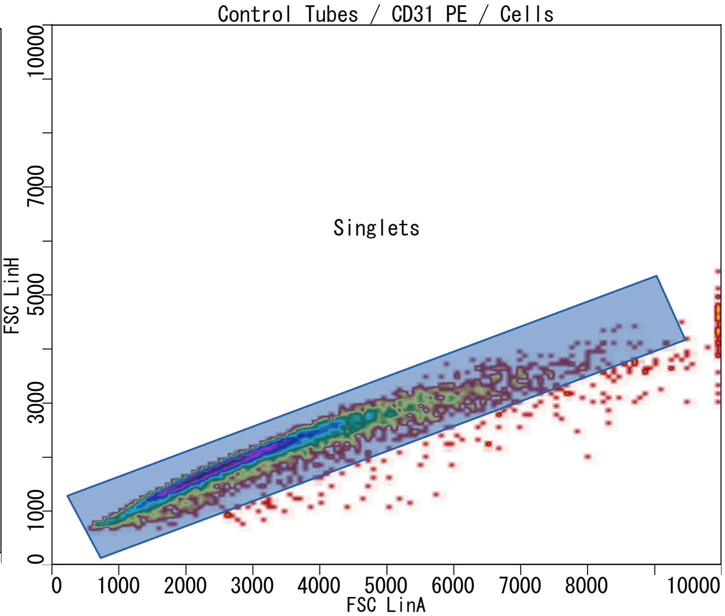


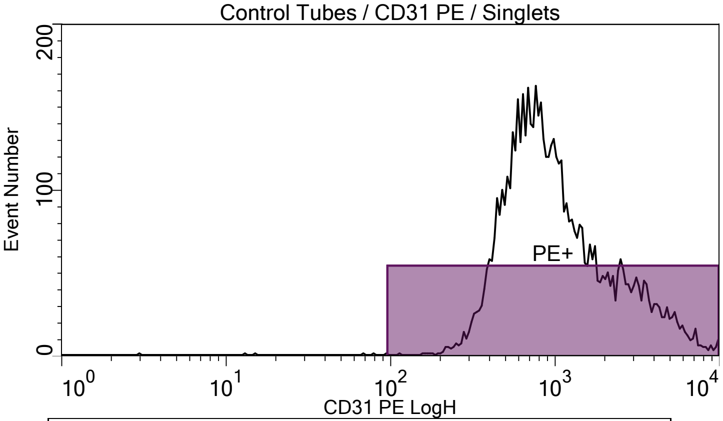


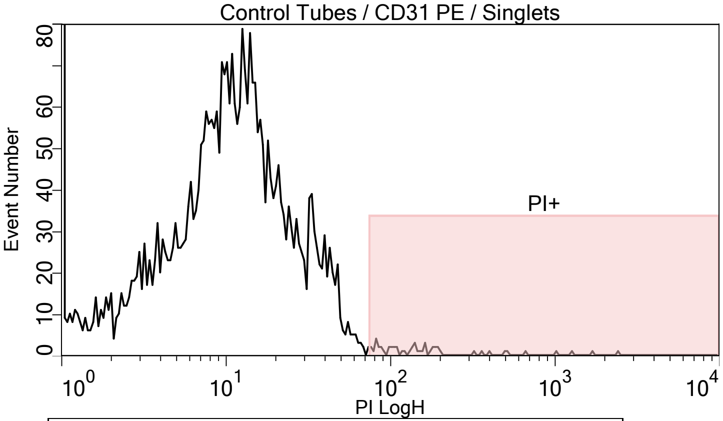


Plot title: Control Tubes / CD31 PE / Singlets
X Axis: CD31 PE LogH

| Population | Event# | % of Parent |
|---|---|---|
| All of Plot | 6095.00 | 100.00 |
| PE+ | 6089.00 | 99.90 |

Plot title: Control Tubes / CD31 PE / Singlets
X Axis: PI LogH

| Population | Event# | % of Parent |
|---|---|---|
| All of Plot | 6095.00 | 100.00 |
| PI+ | 52.00 | 0.85 |

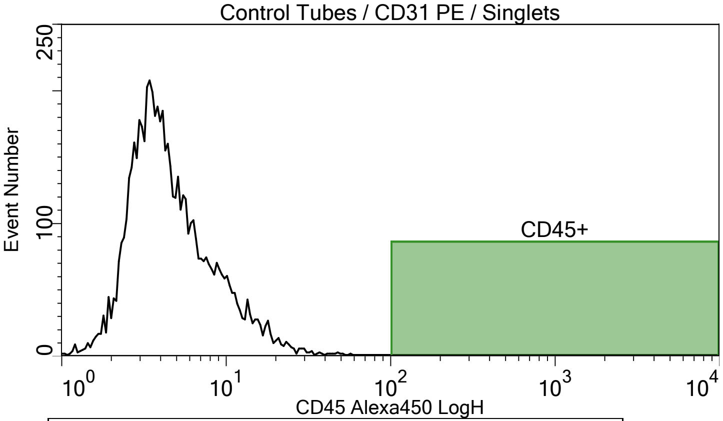


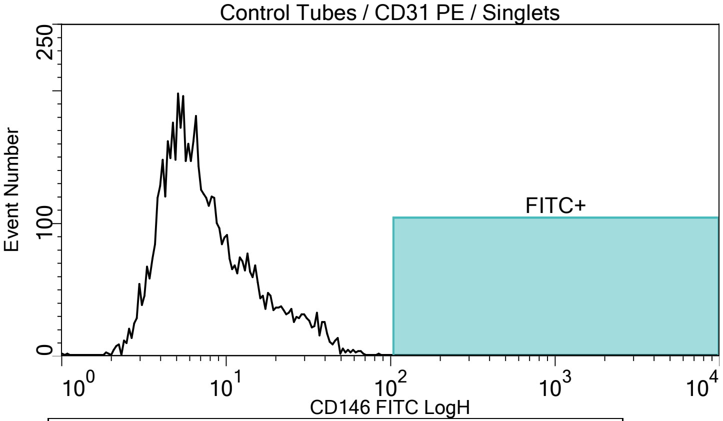


Plot title: Control Tubes / CD31 PE / Singlets
X Axis: CD45 Alexa450 LogH

| Population | Event# | % of Parent |
|---|---|---|
| All of Plot | 6095.00 | 100.00 |
| CD45+ | 0.00 | 0.00 |

Plot title: Control Tubes / CD31 PE / Singlets
X Axis: CD146 FITC LogH

| Population | Event# | % of Parent |
|---|---|---|
| All of Plot | 6095.00 | 100.00 |
| FITC+ | 0.00 | 0.00 |

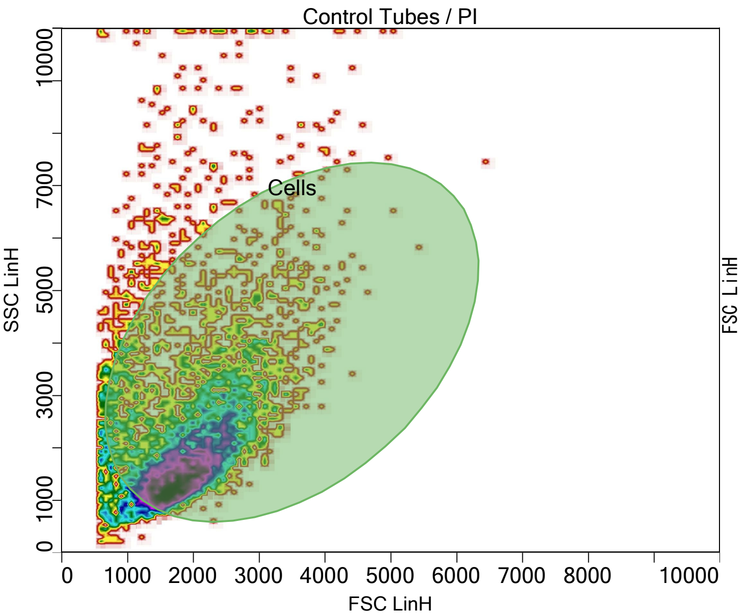


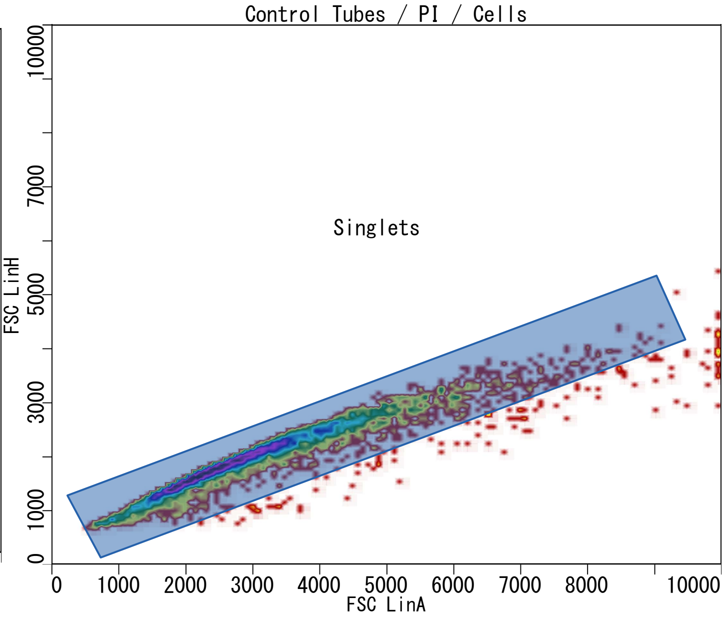


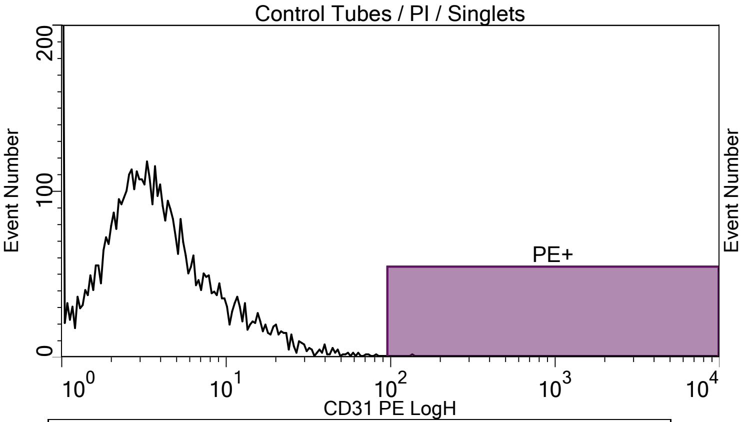


| Plot title: Control Tubes / PI / Singlets<br>X Axis: CD31 PE LogH | | |
|---|---|---|
| **Population** | **Event#** | **% of Parent** |
| All of Plot | 5179.00 | 100.00 |
| PE+ | 1.00 | 0.02 |

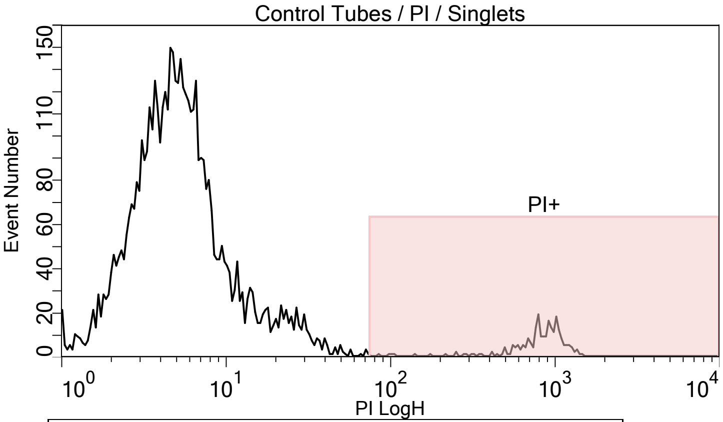


| Plot title: Control Tubes / PI / Singlets<br>X Axis: PI LogH | | |
|---|---|---|
| **Population** | **Event#** | **% of Parent** |
| All of Plot | 5179.00 | 100.00 |
| PI+ | 232.00 | 4.48 |

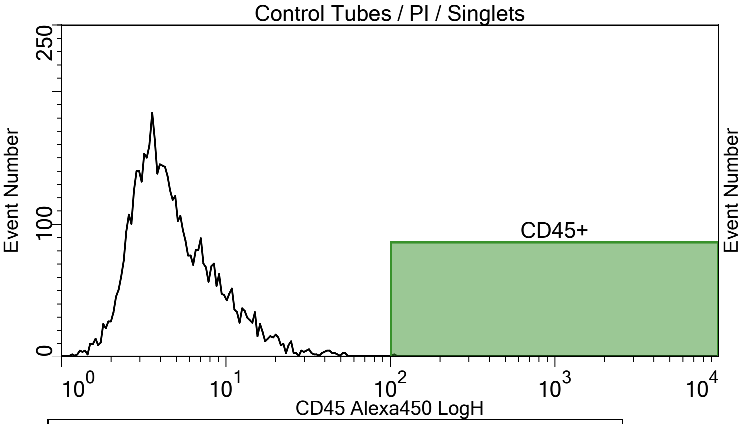


| Plot title: Control Tubes / PI / Singlets<br>X Axis: CD45 Alexa450 LogH | | |
|---|---|---|
| **Population** | **Event#** | **% of Parent** |
| All of Plot | 5179.00 | 100.00 |
| CD45+ | 1.00 | 0.02 |

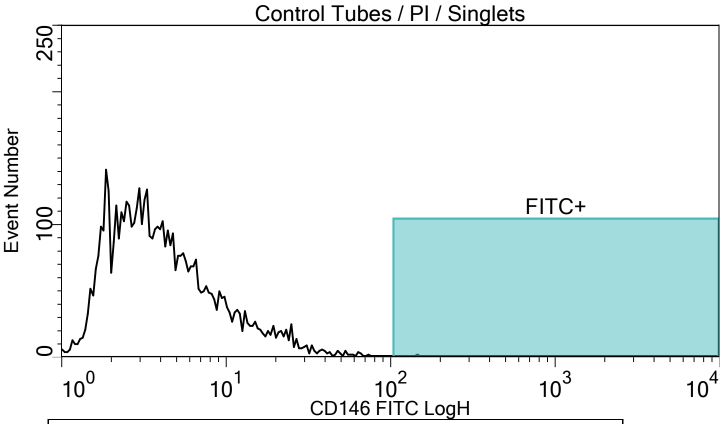


| Plot title: Control Tubes / PI / Singlets<br>X Axis: CD146 FITC LogH | | |
|---|---|---|
| **Population** | **Event#** | **% of Parent** |
| All of Plot | 5179.00 | 100.00 |
| FITC+ | 1.00 | 0.02 |
