## Supplemental Figure 3 for "Pilot Study to Obtain Pulmonary Endothelium from Pediatric Cardiac Catheterization"

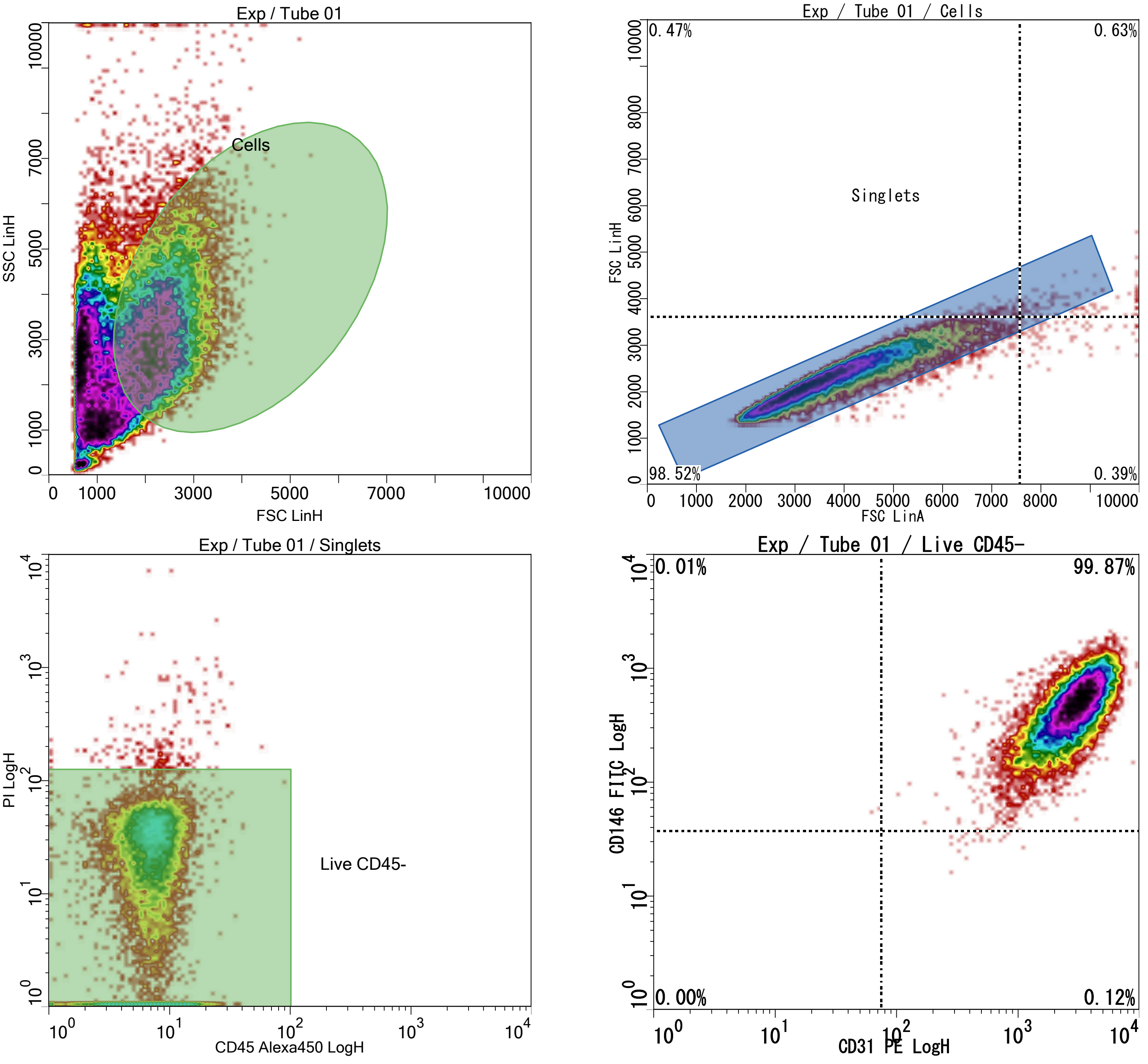


Stratedigm
Cytometry Without Compromise
www.stratedigm.com

Fitc PE PI a450 7-21-25 #21, printed at 7/21/2025 14:52:01
Printed by CellCapTure v4.1 RC27, Copyright 2022 Stratedigm
Licensed to: Oklahoma University OUHSC

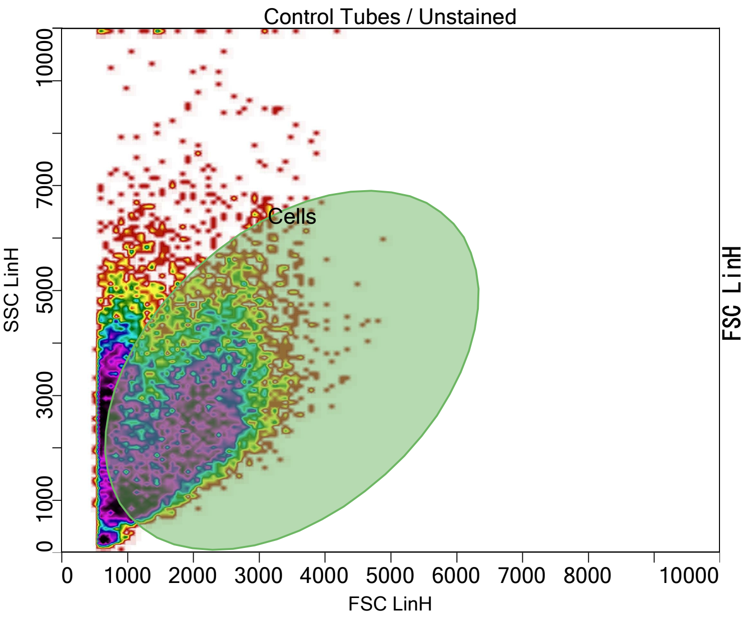


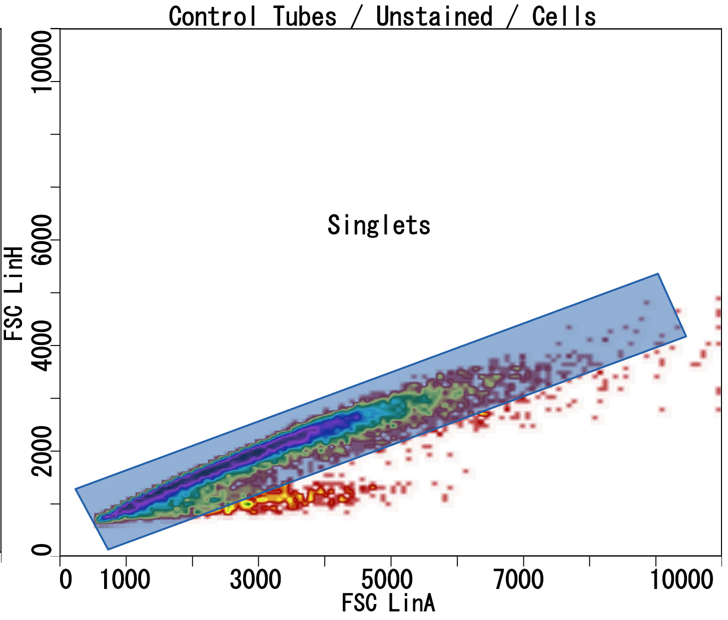


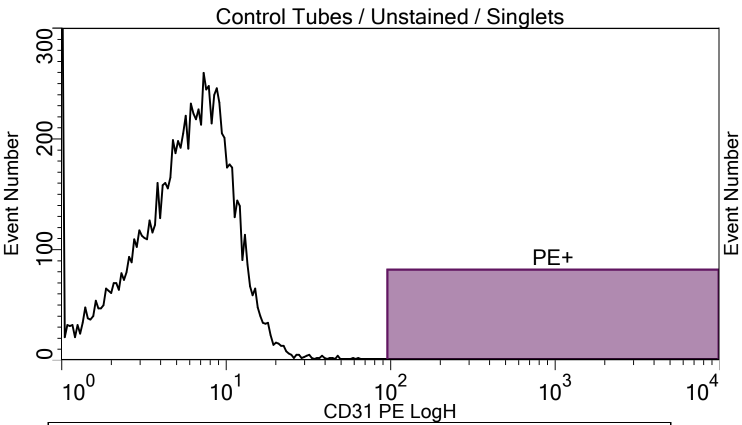


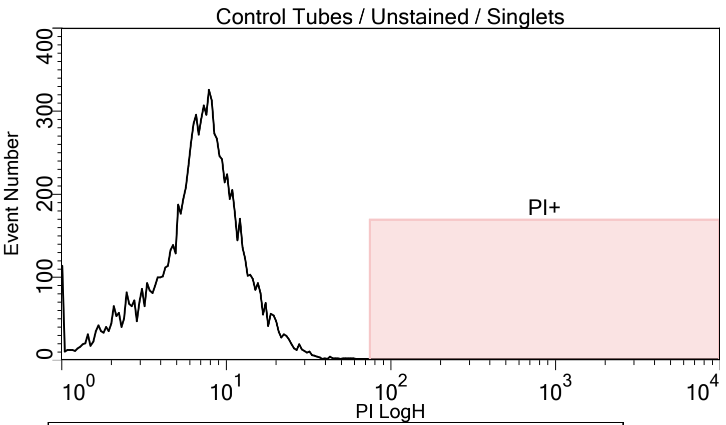


Plot title: Control Tubes / Unstained / Singlets
X Axis: CD31 PE LogH

| Population | Event# | % of Parent |
|---|---|---|
| All of Plot | 10023.00 | 100.00 |
| PE+ | 0.00 | 0.00 |

Plot title: Control Tubes / Unstained / Singlets
X Axis: PI LogH

| Population | Event# | % of Parent |
|---|---|---|
| All of Plot | 10023.00 | 100.00 |
| PI+ | 0.00 | 0.00 |

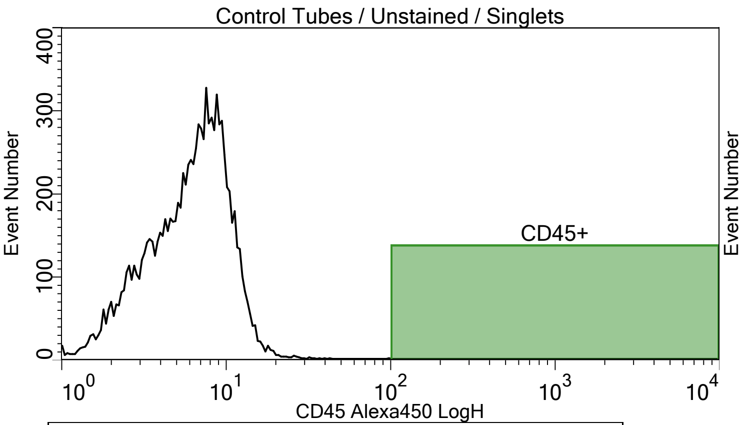


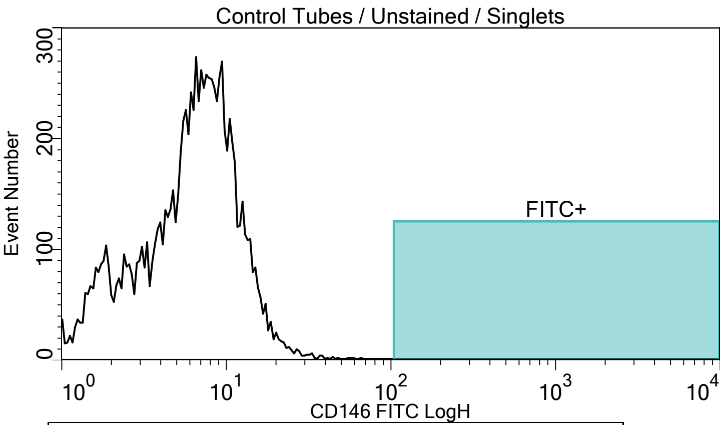


Plot title: Control Tubes / Unstained / Singlets
X Axis: CD45 Alexa450 LogH

| Population | Event# | % of Parent |
|---|---|---|
| All of Plot | 10023.00 | 100.00 |
| CD45+ | 0.00 | 0.00 |

Plot title: Control Tubes / Unstained / Singlets
X Axis: CD146 FITC LogH

| Population | Event# | % of Parent |
|---|---|---|
| All of Plot | 10023.00 | 100.00 |
| FITC+ | 0.00 | 0.00 |

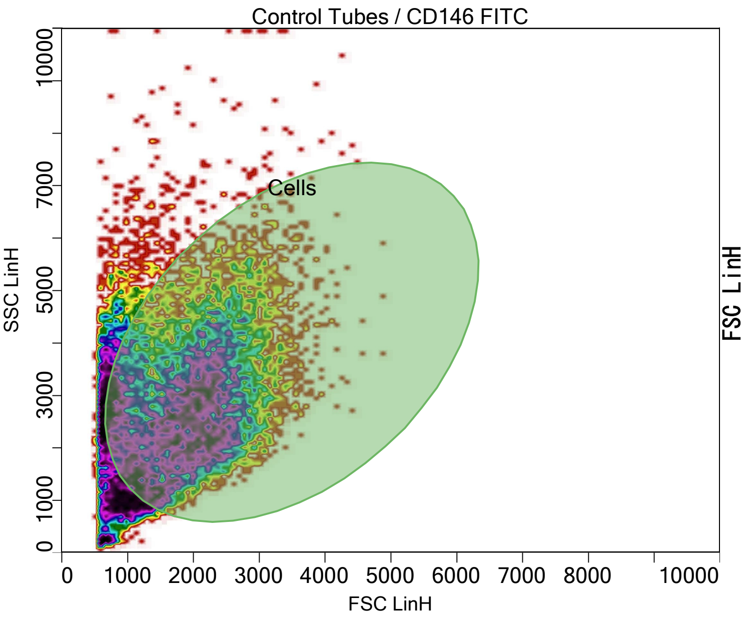


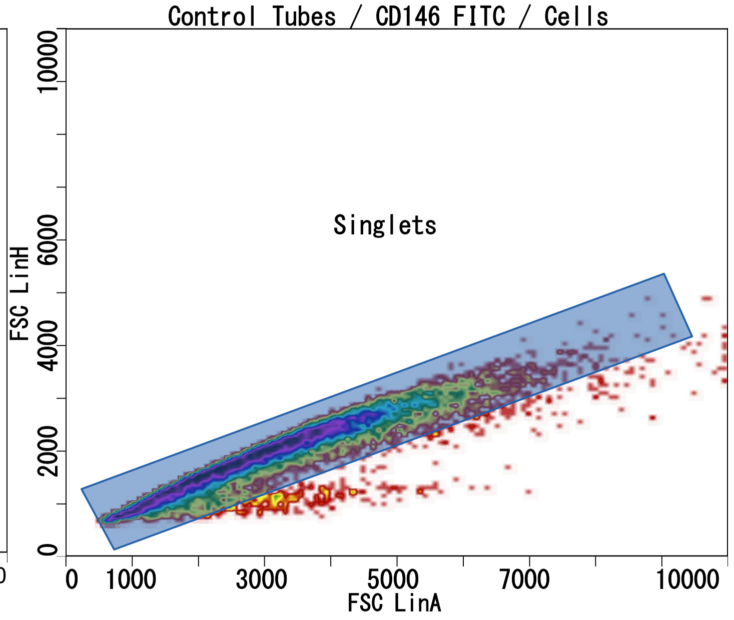


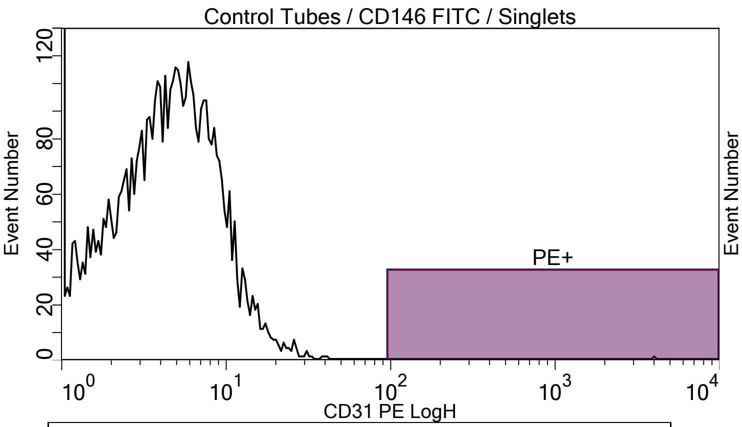


Plot title: Control Tubes / CD146 FITC / Singlets
X Axis: CD31 PE LogH

| Population | Event# | % of Parent |
|---|---|---|
| All of Plot | 10037.00 | 100.00 |
| PE+ | 1.00 | 0.01 |

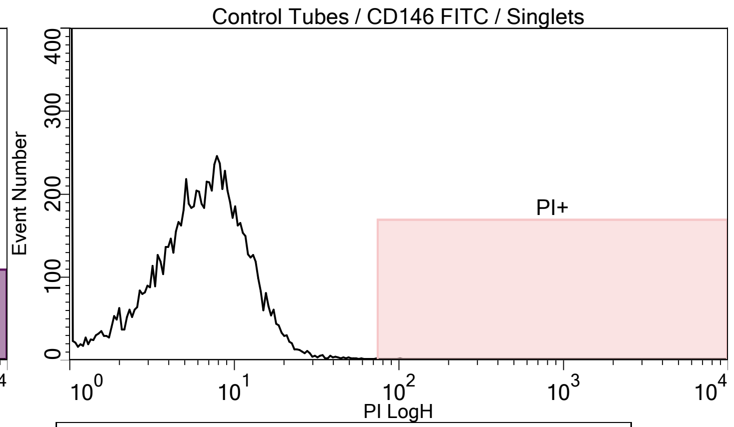


Plot title: Control Tubes / CD146 FITC / Singlets
X Axis: PI LogH

| Population | Event# | % of Parent |
|---|---|---|
| All of Plot | 10037.00 | 100.00 |
| PI+ | 1.00 | 0.01 |

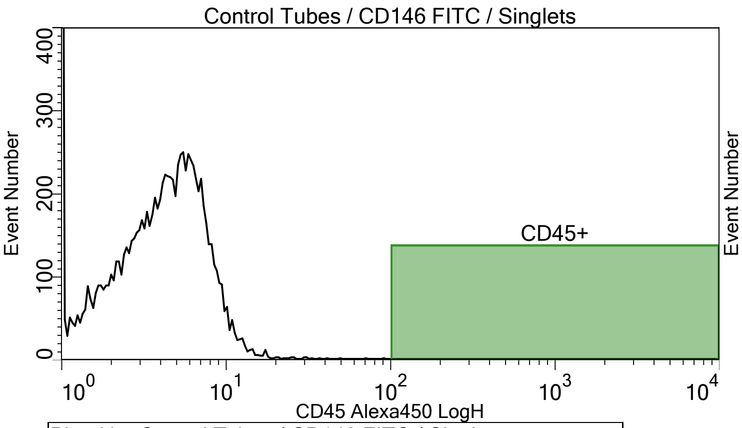


Plot title: Control Tubes / CD146 FITC / Singlets
X Axis: CD45 Alexa450 LogH

| Population | Event# | % of Parent |
|---|---|---|
| All of Plot | 10037.00 | 100.00 |
| CD45+ | 0.00 | 0.00 |

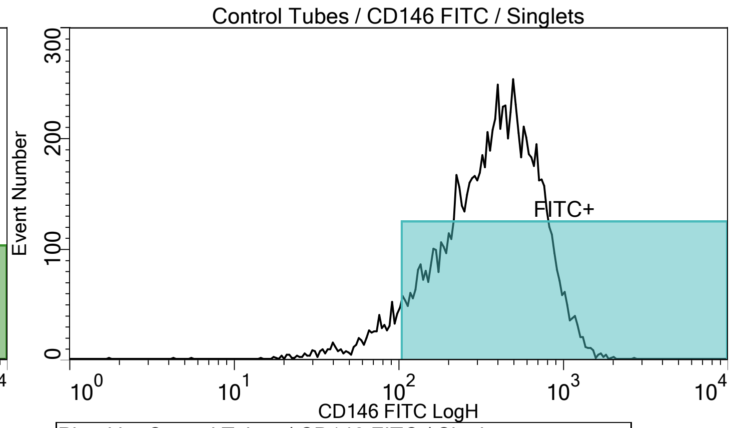


Plot title: Control Tubes / CD146 FITC / Singlets
X Axis: CD146 FITC LogH

| Population | Event# | % of Parent |
|---|---|---|
| All of Plot | 10037.00 | 100.00 |
| FITC+ | 9348.00 | 93.14 |

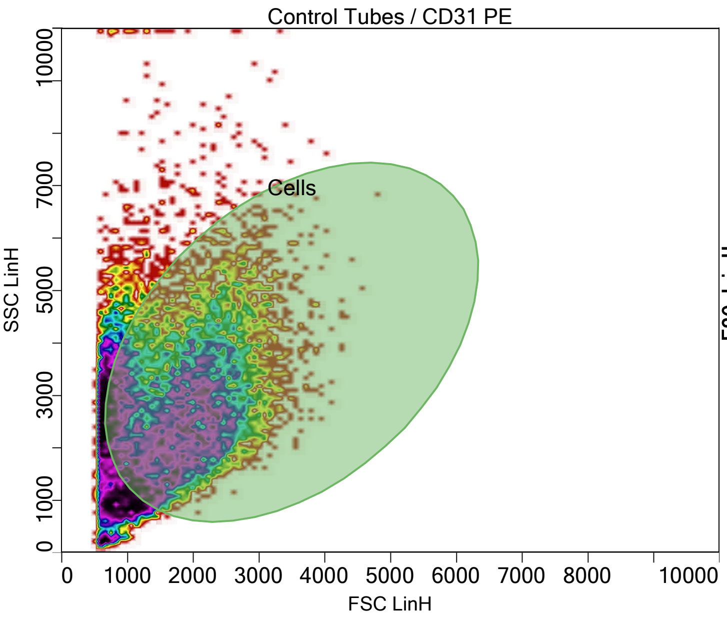


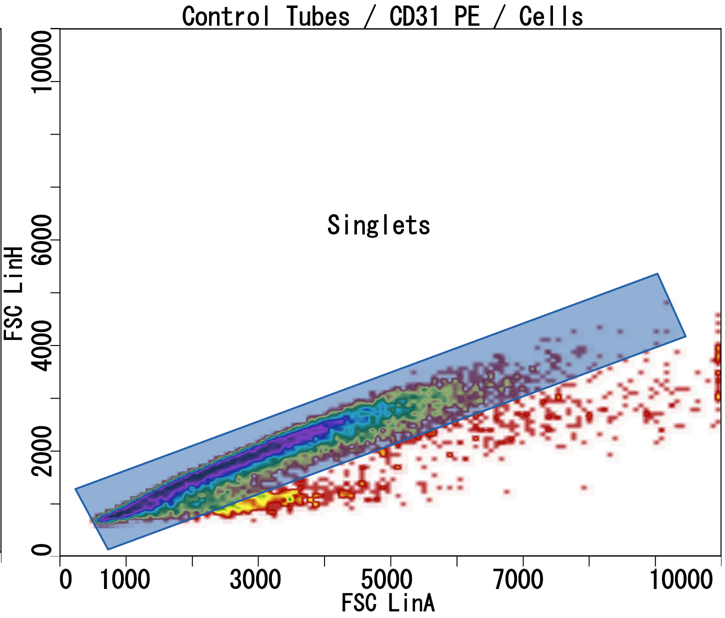


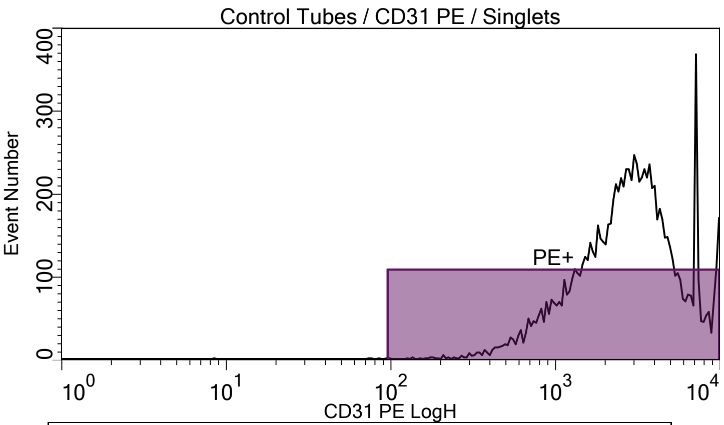


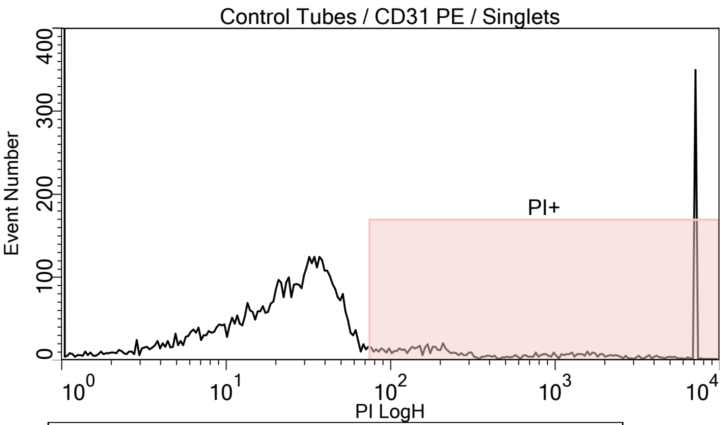


Plot title: Control Tubes / CD31 PE / Singlets
X Axis: CD31 PE LogH

| Population | Event# | % of Parent |
|---|---|---|
| All of Plot | 10050.00 | 100.00 |
| PE+ | 10045.00 | 99.95 |

Plot title: Control Tubes / CD31 PE / Singlets
X Axis: PI LogH

| Population | Event# | % of Parent |
|---|---|---|
| All of Plot | 10050.00 | 100.00 |
| PI+ | 1075.00 | 10.70 |

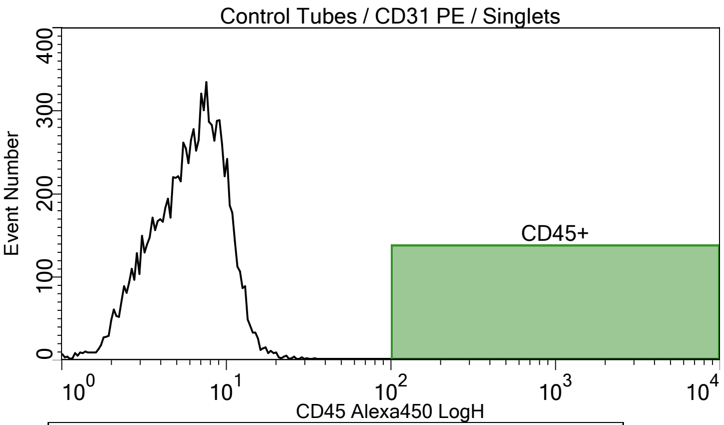


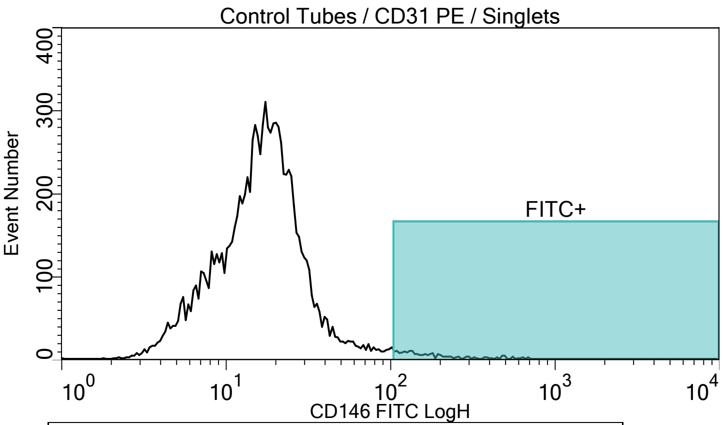


Plot title: Control Tubes / CD31 PE / Singlets
X Axis: CD45 Alexa450 LogH

| Population | Event# | % of Parent |
|---|---|---|
| All of Plot | 10050.00 | 100.00 |
| CD45+ | 0.00 | 0.00 |

Plot title: Control Tubes / CD31 PE / Singlets
X Axis: CD146 FITC LogH

| Population | Event# | % of Parent |
|---|---|---|
| All of Plot | 10050.00 | 100.00 |
| FITC+ | 182.00 | 1.81 |

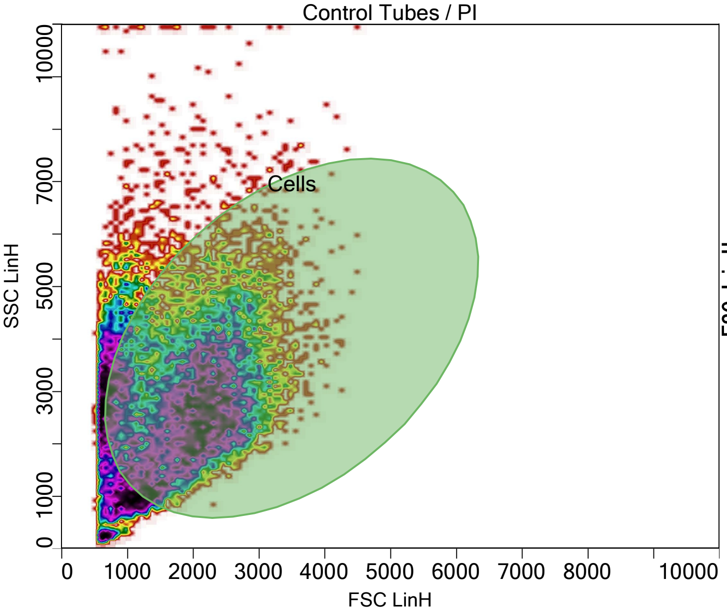


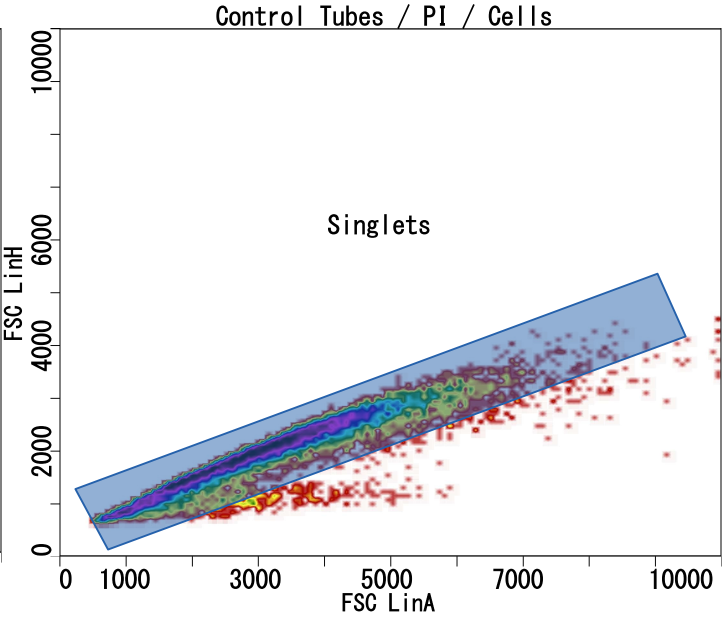


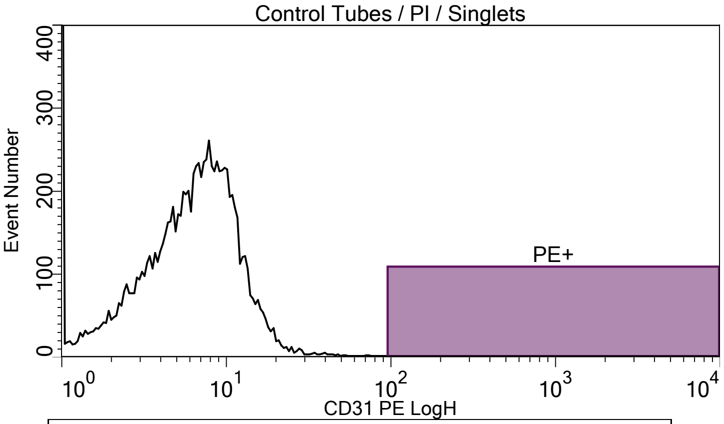


| Plot title: Control Tubes / PI / Singlets | | |
|---|---|---|
| X Axis: CD31 PE LogH | | |
| **Population** | **Event#** | **% of Parent** |
| All of Plot | 10029.00 | 100.00 |
| PE+ | 0.00 | 0.00 |

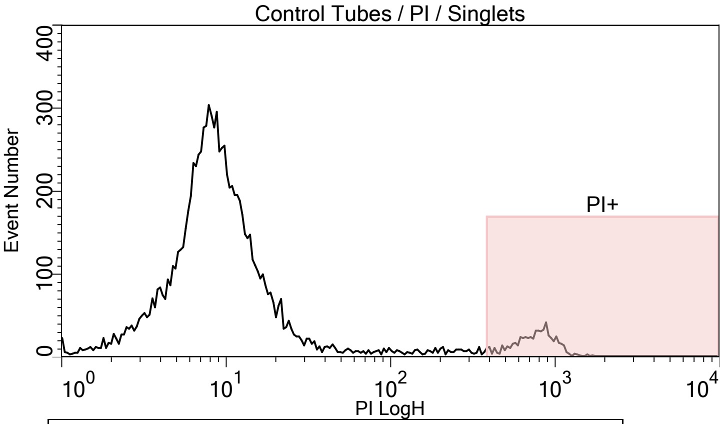


| Plot title: Control Tubes / PI / Singlets | | |
|---|---|---|
| X Axis: PI LogH | | |
| **Population** | **Event#** | **% of Parent** |
| All of Plot | 10029.00 | 100.00 |
| PI+ | 569.00 | 5.67 |

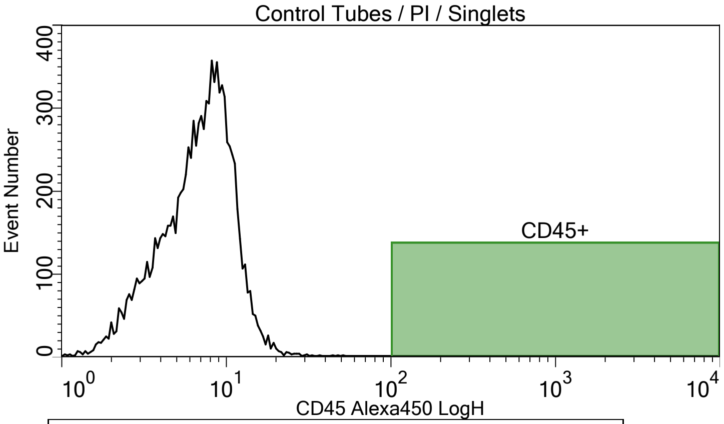


| Plot title: Control Tubes / PI / Singlets | | |
|---|---|---|
| X Axis: CD45 Alexa450 LogH | | |
| **Population** | **Event#** | **% of Parent** |
| All of Plot | 10029.00 | 100.00 |
| CD45+ | 0.00 | 0.00 |

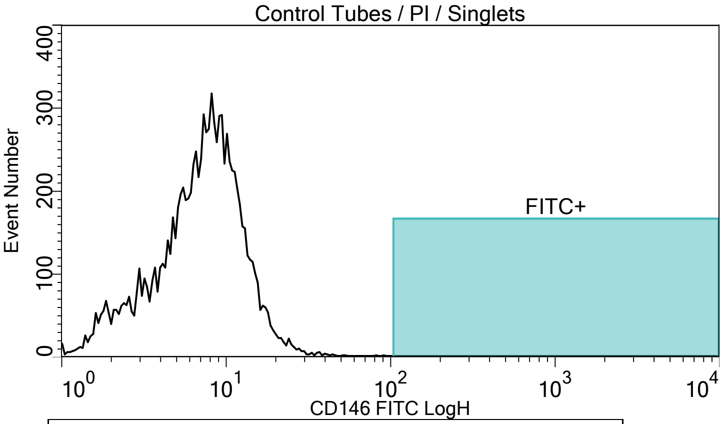


| Plot title: Control Tubes / PI / Singlets | | |
|---|---|---|
| X Axis: CD146 FITC LogH | | |
| **Population** | **Event#** | **% of Parent** |
| All of Plot | 10029.00 | 100.00 |
| FITC+ | 0.00 | 0.00 |

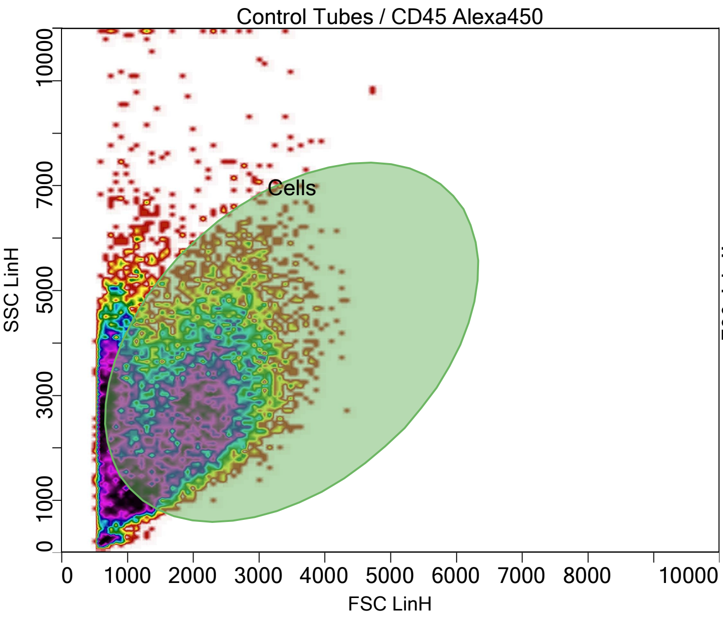


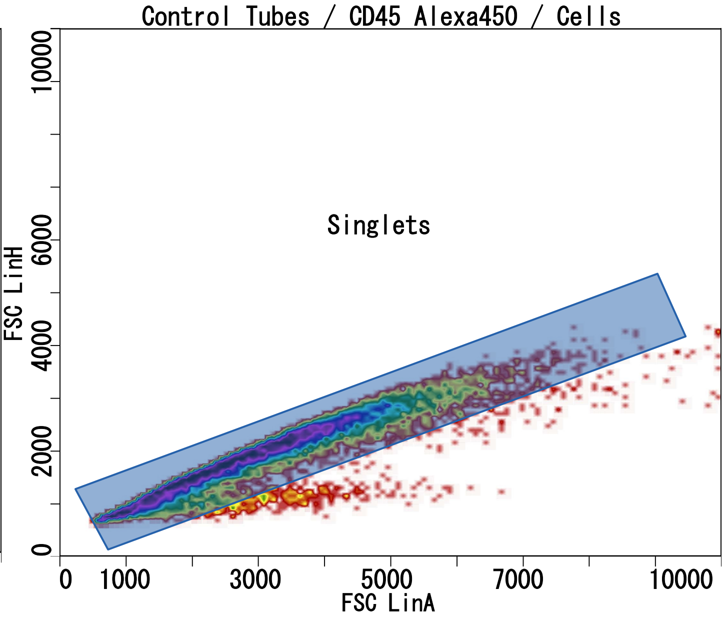


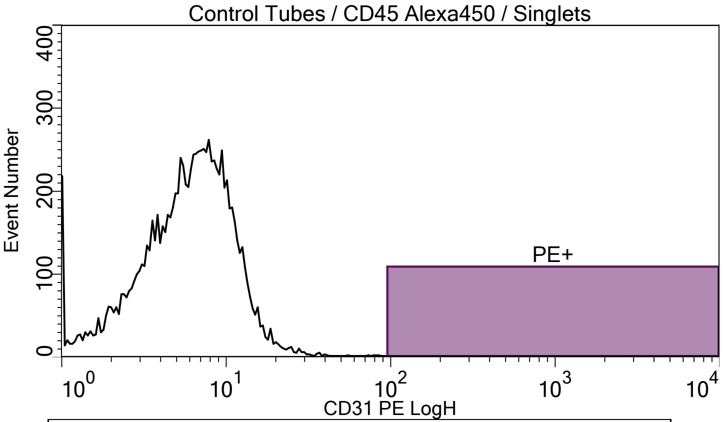


Plot title: Control Tubes / CD45 Alexa450 / Singlets
X Axis: CD31 PE LogH

| Population | Event# | % of Parent |
|---|---|---|
| All of Plot | 10018.00 | 100.00 |
| PE+ | 0.00 | 0.00 |

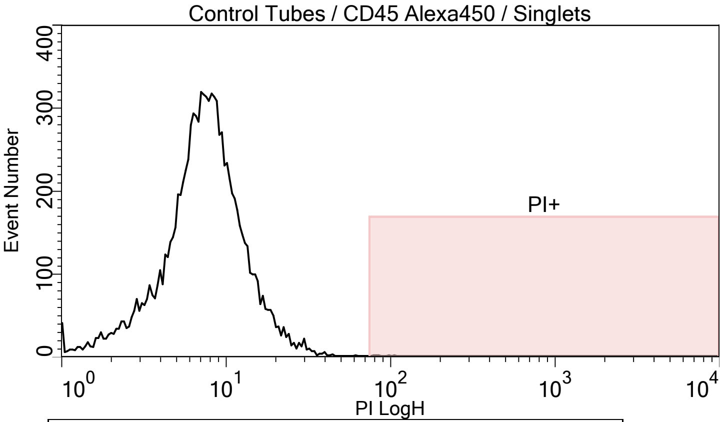


Plot title: Control Tubes / CD45 Alexa450 / Singlets
X Axis: PI LogH

| Population | Event# | % of Parent |
|---|---|---|
| All of Plot | 10018.00 | 100.00 |
| PI+ | 5.00 | 0.05 |

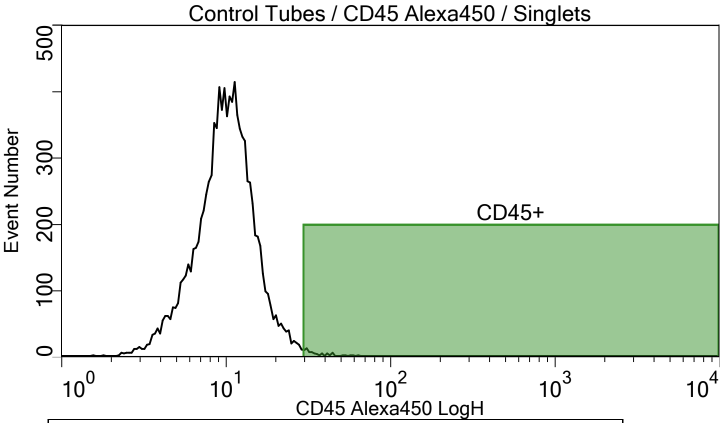


Plot title: Control Tubes / CD45 Alexa450 / Singlets
X Axis: CD45 Alexa450 LogH

| Population | Event# | % of Parent |
|---|---|---|
| All of Plot | 10018.00 | 100.00 |
| CD45+ | 60.00 | 0.60 |

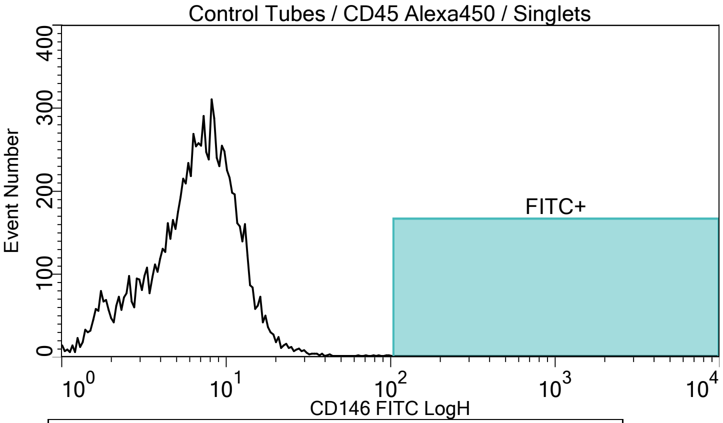


Plot title: Control Tubes / CD45 Alexa450 / Singlets
X Axis: CD146 FITC LogH

| Population | Event# | % of Parent |
|---|---|---|
| All of Plot | 10018.00 | 100.00 |
| FITC+ | 0.00 | 0.00 |
