## Supplementary figures and images for "Pilot Study to Obtain Pulmonary Endothelium from Pediatric Cardiac Catheterization"

### Supplemental Figure 4

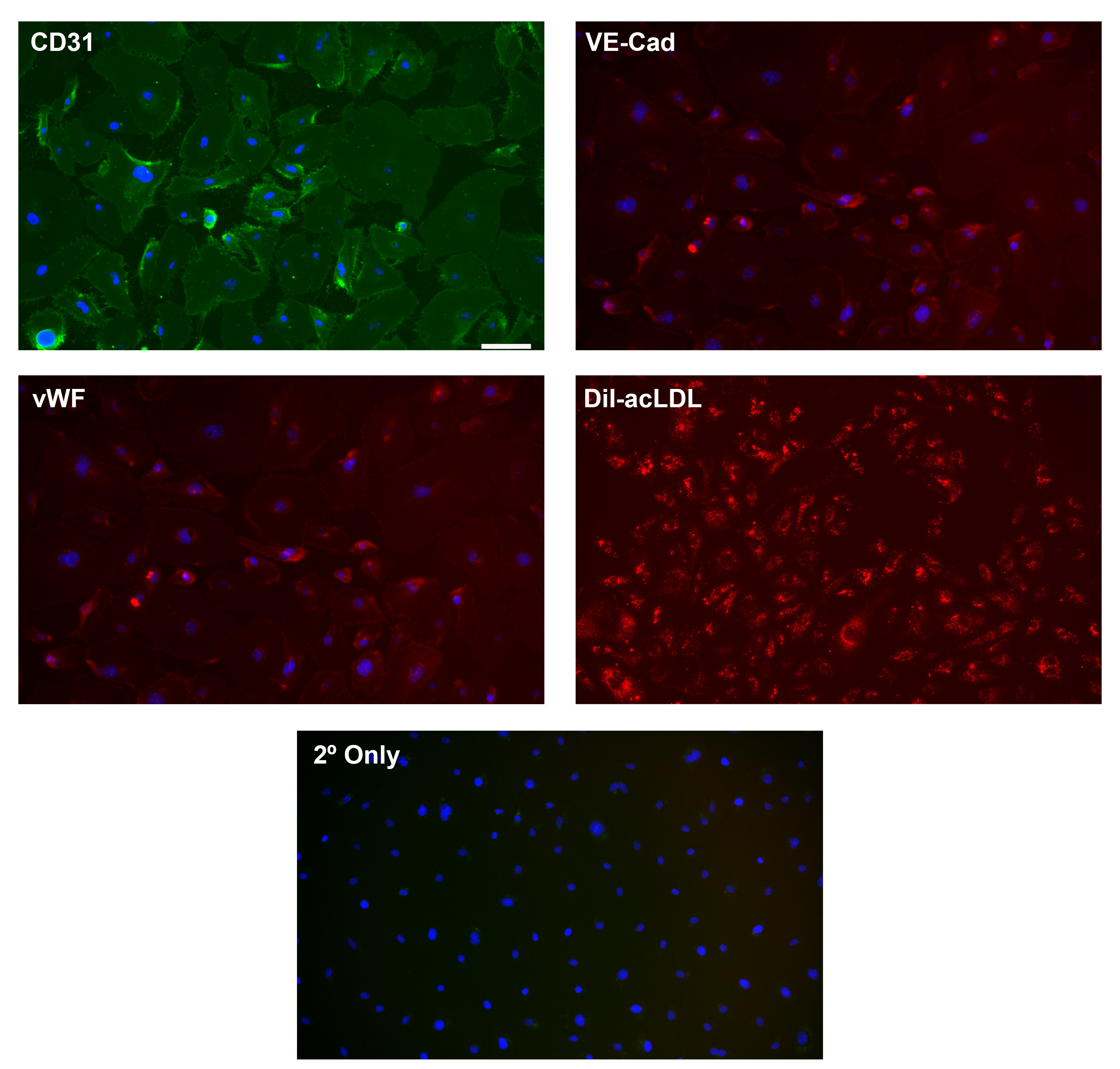

### Supplemental Figure 5

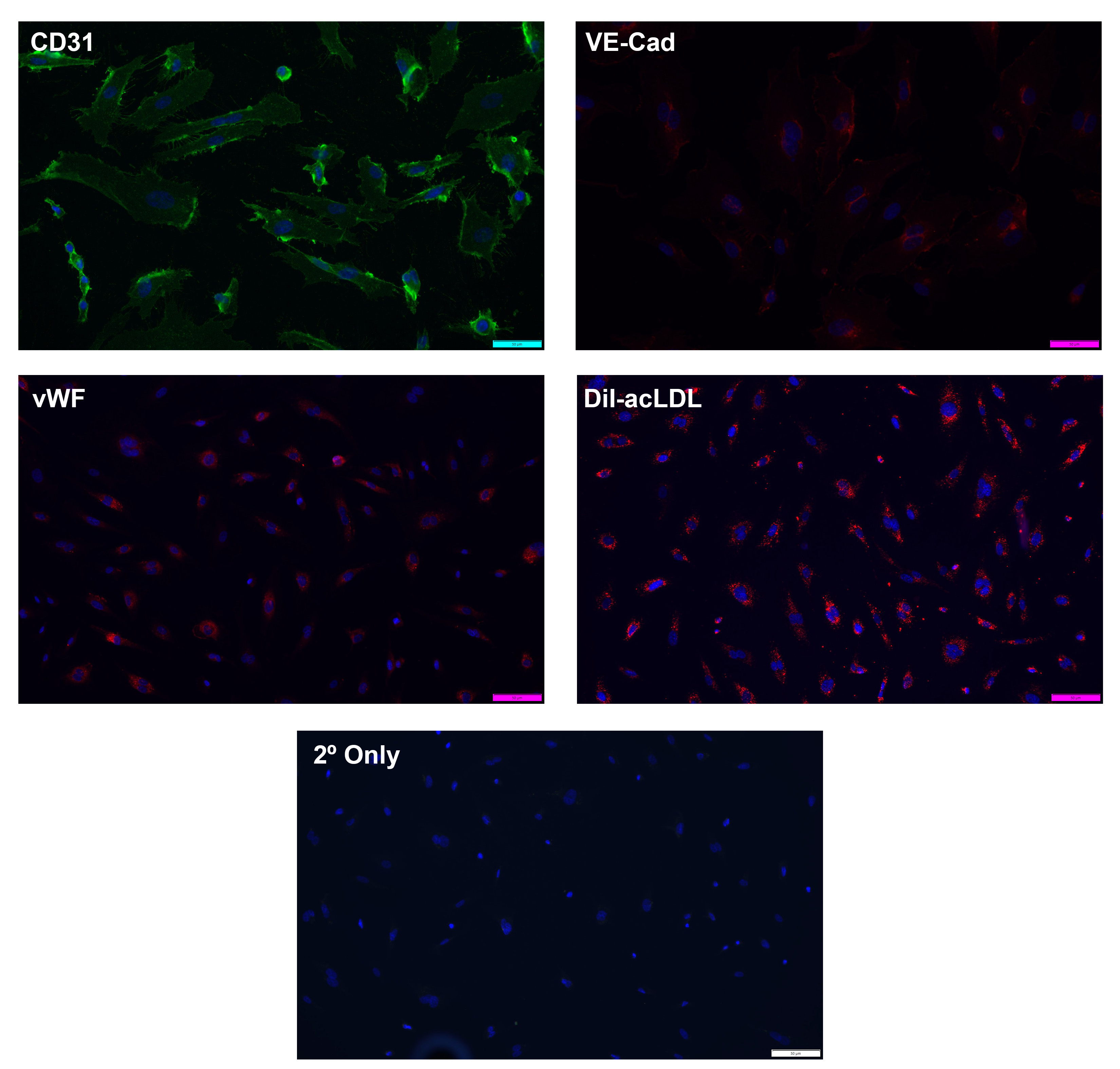
