## Supplemental Legend for "Pilot Study to Obtain Pulmonary Endothelium from Pediatric Cardiac Catheterization"

**Figure S1:** Flow cytometry controls and gating for subject 17.

**Figure S2:** Flow cytometry controls and gating for subject 19.

**Figure S3:** Flow cytometry controls and gating for subject 21.

**Figure S4, S5:** Representative images of formalin fixed cells from subject 17 staining for endothelial markers CD31, vascular endothelial cadherin (VE-Cad), von Willebrand Factor (vWF), and 4 hour uptake of DiI-conjugated acetylated LDL (DiI-acLDL). All images obtained at 20x magnification, scale bars are 50μm. Images obtained in grayscale and converted to pseudocolor in ImageJ.
